# DNA repair biomarkers to guide usage of combined PARP inhibitors and chemotherapy: a meta-analysis and systematic review

**DOI:** 10.1101/2023.05.24.23290442

**Authors:** Zoe Phan, Caroline E. Ford, C. Elizabeth Caldon

**Affiliations:** The Kinghorn Cancer Centre, Garvan Institute of Medical Research, Sydney, NSW, 2010, Australia; St. Vincent’s Clinical School, Faculty of Medicine, University of New South Wales, Sydney, NSW, 2052, Australia; School of Clinical Medicine, Faculty of Medicine and Health, University of New South Wales, Sydney, NSW, 2052, Australia

**Keywords:** PARP inhibitor, homologous recombination deficiency, chemotherapy, combination therapy, BRCA1/2, SFLN11, survival, DNA repair

## Abstract

**Purpose:** The addition of PARP inhibitors to chemotherapy has been assessed in ∼80 clinical trials across multiple malignancies, on the premise that PARP inhibitors will increase chemotherapy effectiveness regardless of whether cancers have underlying disruption of DNA repair pathways. Consequently, the majority of combination therapy trials have been performed on patients without biomarker selection, despite the use of homologous recombination deficiency to dictate use of PARP inhibitors in the maintenance setting. An unresolved question is whether biomarkers are needed to identify patients who respond to combination PARP inhibitors and chemotherapy.

**Methods:** A systematic literature review identified studies using PARP inhibitors in combination with chemotherapy versus chemotherapy alone, where the study included a biomarker of DNA repair function (*BRCA1*, *BRCA2*, BRCAPRO, ATM, ERCC1, SFLN11). Hazard ratios (HR) were pooled in a meta-analysis using generic inverse-variance and fixed or random effects modelling. Subgroup analyses were conducted on biomarker selection and type of malignancy.

**Results:** Nine studies comprising 2,084 patients met the inclusion criteria. Progression-free survival (PFS) was significantly better in patients with a DNA repair biomarker (HR 0.52, 95% confidence interval (CI) 0.43-0.63; p < 0.00001), but there was no benefit in patients who lacked a biomarker (HR 0.94, 95% CI 0.82–1.08; p = 0.38). Subgroup analysis showed that *BRCA* mutation and SFLN11 biomarkers could predict benefit, and biomarker-driven benefit occurred in ovarian, breast and small cell lung cancers. The addition of PARP inhibitors was associated with increased grade 3/4 side effects, and particularly neutropenia.

**Conclusions:** Combination therapy only increases PFS in patients with identifiable DNA repair biomarkers. This indicates that PARP inhibitors do not sensitise patients to chemotherapy treatment, except where their cancer has a homologous recombination defect, or an alternative biomarker of altered DNA repair. While effective in patients with DNA repair biomarkers, there is a risk of high-grade haematological side-effects with the use of combination therapy. Thus, the benefit in PFS from combination therapy must be weighed against potential adverse effects, as individual arms of treatment can also confer benefit.

**GRAPHICAL ABSTRACT:** 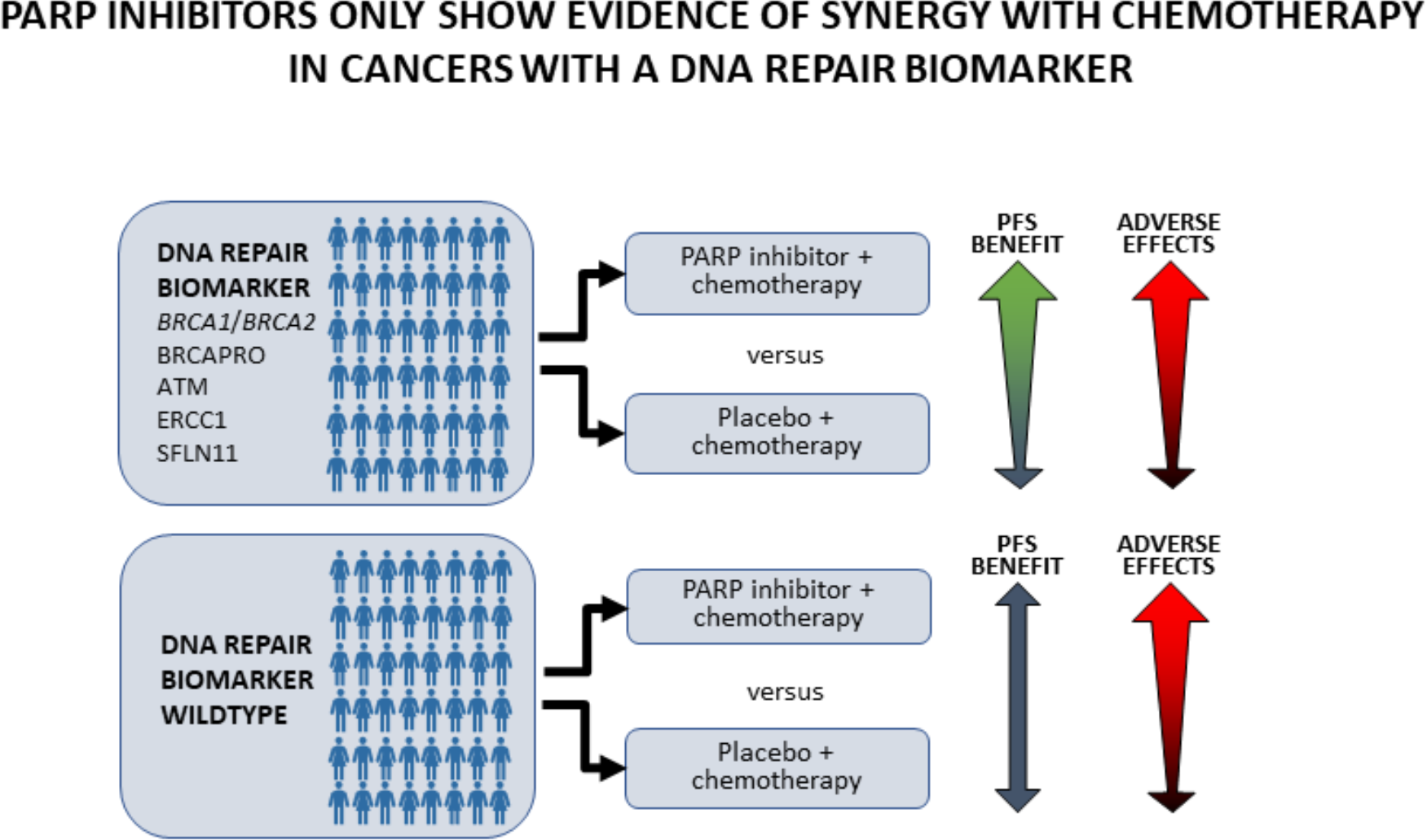

## INTRODUCTION

Cancer places an enormous burden on our healthcare system as the leading cause of death worldwide, accounting for nearly 10 million deaths in 2020 [1]. Targeted therapies and personalised medicines are now demonstrating significant benefit in reducing cancer-associated mortality [2]. Poly (ADP-ribose) polymerase (PARP)-inhibitors were introduced as a targeted therapy in 2014, with initial approvals to treat cancers that had defects in homologous recombination, such as mutations in the genes *BRCA1* and *BRCA2*. Alterations to homologous recombination-related genes lead to a homologous recombination deficiency where cells are unable to accurately repair DNA breaks and become overly reliant on other DNA repair pathways, and particularly PARP-mediated repair. PARP inhibitors have also been investigated in cancers with alterations in other DNA repair related genes [3]. When PARP is inhibited or trapped on DNA in a cell with DNA repair deficiency, these cells rely on more error-prone pathways of repair, and this leads to the accumulation of DNA damage and subsequent cell death (**Figure 1A)**.

**Figure 1.**
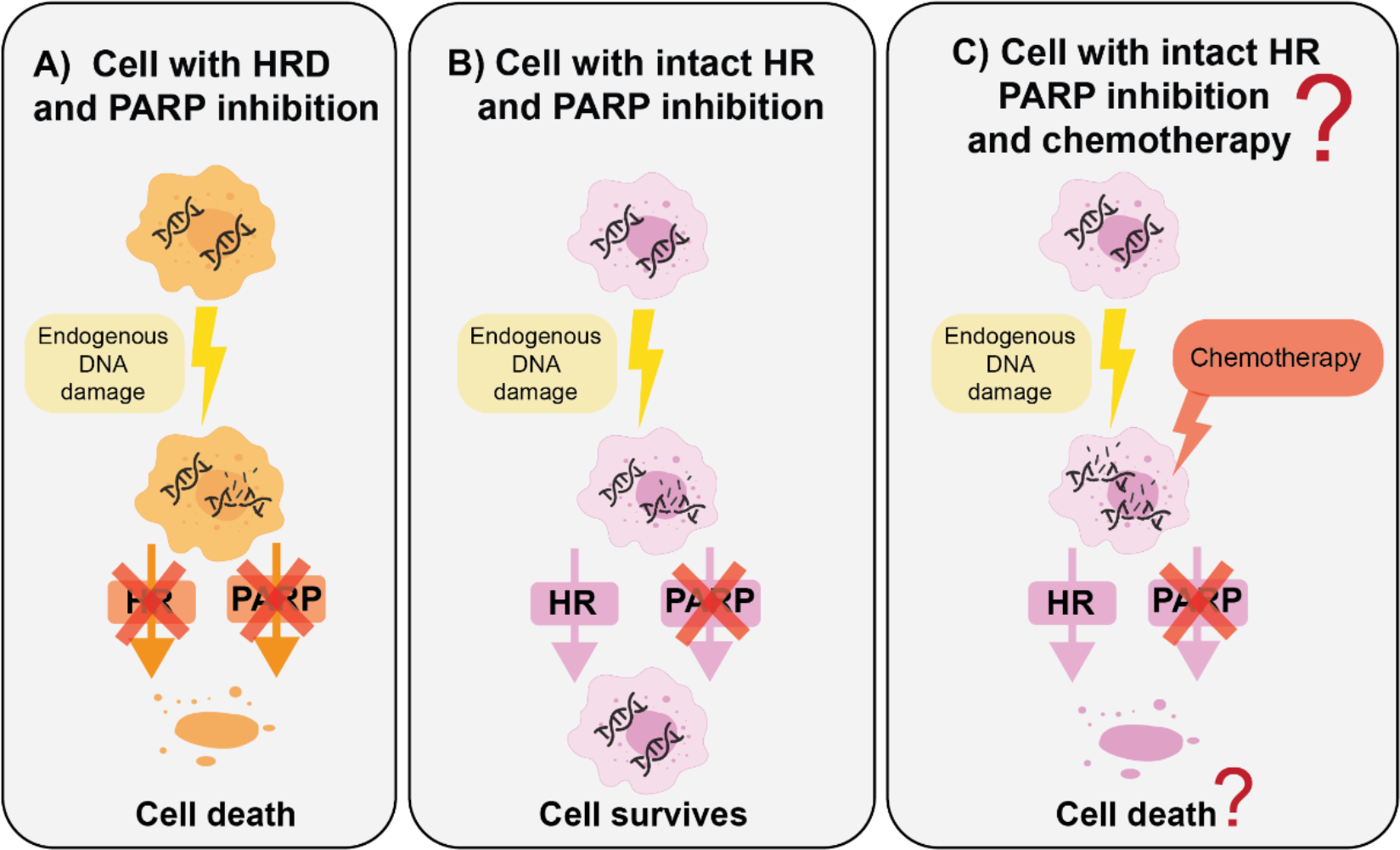
Schematic of how chemotherapy could potentiate PARP inhibition. In Panel A, a cancer cell with homologous recombination deficiency (HRD) is sensitive to PARP inhibitors because cells are unable to properly repair low level DNA damage, leading to cancer cell death. Low level DNA damage is constantly generated by common environmental exposures (UV, radiation) and endogenous DNA damage from oxidation and DNA metabolism [4]. In Panel B, a cell with intact homologous recombination (HR) is able to repair low level damage through homologous recombination, even in the presence of PARP inhibitors. In Panel C, a cell with intact homologous recombination may be sensitised to chemotherapy by inhibiting DNA repair with PARP inhibitors.

Four PARP inhibitors are approved for clinical use as monotherapies: olaparib, rucaparib, niraparib and talazoparib. Olaparib was first approved as a maintenance therapy for patients with platinum-sensitive recurrent germline *BRCA*-mutated ovarian cancer who had a complete or partial response to platinum-based chemotherapy [5]. Olaparib has since been approved for HER2-negative, germline *BRCA-*mutated breast cancer, and a subset of pancreatic cancer patients [6, 7]. In 2016, rucaparib was approved as a monotherapy for patients with advanced ovarian cancer with either somatic or germline *BRCA* mutations [8]. In 2018, talazoparib was approved for the treatment of deleterious germline *BRCA*-mutated, HER2-negative metastatic breast cancer [9]. Niraparib has demonstrated benefit in ovarian cancer patients who have previously responded to platinum chemotherapy, regardless of their homologous recombination deficiency status [8, 10, 11] which has led to its approval in the maintenance setting for newly diagnosed and recurrent ovarian cancers with platinum response. Additional PARP inhibitors, such as veliparib, are currently being evaluated in clinical trials [12–16].

A critical aspect of PARP inhibitor usage is to accurately identify patients that will benefit from these targeted therapies. The most common predictive biomarkers to indicate PARP inhibitor sensitivity are mutations in the genes, *BRCA1* and *BRCA2,* which are essential in maintaining genomic integrity. Identifying pathogenic *BRCA1*/*BRCA2* mutations can be further refined by using genetic tests such as Myriad BRACAnalysis CDx, to determine germline *BRCA1/BRCA2* mutations, or statistical models such as BRCAPRO, which incorporate family and individual history of cancer diagnoses. Homologous recombination deficiency can also be driven by other genomic changes including *BRCA1* methylation, or *PALB2*/*RAD51C*/*RAD51D* mutations [17]. However, these aberrations are not yet used clinically to identify patients for treatment with PARP inhibitors. The presence of these mutations can give rise to characteristic genomic rearrangements indicative of the loss of homologous recombination, which is often termed “genomic scarring”. Genomic scars include loss of heterozygosity, large-scale translocations and telomeric allelic imbalance. Next-generation sequencing-based assays such as myChoice CDx can be used as an alternative or adjunct test to detect these genomic rearrangements and identify homologous recombination deficient (HRD) patients within the clinic, and this approach is commonly referred to as ‘HRD-score’ (**Table 1**).

**Table 1.** Markers of DNA repair deficiency.

| Biomarker | Method of measurement | Cut-off / Discriminating factor |
| --- | --- | --- |
| *BRCA* | DNA | Documented deleterious *BRCA1/2* mutation, Myriad BRACAnalysis CDx or BRCAPRO score of > 30% (this is an algorithm that predicts whether a person has inherited a *BRCA* gene mutation) |
| HRD-score | DNA | HRD-score of > 42 |
| SLFN11 | IHC | H-score $\geq 1$ |
| ATM | IHC | A clinical status of ATM-positive was assigned to cases with 25% or more gastric cancer cells exhibiting nuclear staining (of weak, moderate, or strong staining intensity) and an ATM-negative status to cases with less than 25% nuclear staining. |
| ERCC1 | IHC | Expression of ERCC1 was defined by immunohistochemistry scores as high (2+, 3+) or low (0, 1+) |

HRD-score: Homologous recombination deficiency score.

New markers of altered DNA repair potential are also emerging (e.g. ATM, ERCC1, SLFN11) and being assessed pre-clinically and in clinical trials. ATM is a key activator of the DNA damage response to double-strand breaks and in pre-clinical models, loss/depletion of ATM was shown to increase sensitivity of cells to PARP inhibitors in gastric, colorectal, lung and pancreatic cancer [18]. Clinical trials are now assessing the benefit of PARP inhibitors in ATM-deficient prostate and gastric cancer patients [19, 20]. ERCC1 is a key mediator of nucleotide excision DNA repair, and it has been proposed that PARP inhibition increases sensitivity of ERCC1-low non-small cell lung cancer cells to cisplatin [21]. SLFN11 is implicated in replication fork stress and has been studied in several pre-clinical models of small cell lung cancer where high SLFN11 expression strongly predicted cisplatin and PARP inhibitor response [22–24].

Since the realisation of the clinical benefit of PARP inhibitors, a significant effort had been made to expand their use to treat cancers without underlying DNA repair defects. Thus, a large body of research has investigated whether there is synergy between PARP inhibitors and other therapies, including chemotherapies. The premise for synergy is that excessive DNA damage from chemotherapy will create too many DNA breaks and adducts to be repaired when PARP is simultaneously inhibited, resulting in lethality (**Figure 1B**). This is hypothesised to occur irrespective of defects in DNA repair [25–27].

Multiple preclinical *in vitro* and *in vivo* studies have demonstrated that synergy between PARP inhibitors and chemotherapies can occur in the absence of identifiable DNA repair defects. Several studies that combine PARP inhibitors with DNA alkylating agent temozolimide (TMZ) have reported synergy: with niraparib across *in vitro* wildtype pheochromocytoma models [28], with olaparib across *in vitro* and *in vivo* xenograft models of chordoma [29], with niraparib for multiple myeloma [30], with veliparib in melanoma and glioma mouse models [31], and PARP inhibitor GPI 15427 in combination with TMZ and CPT-11 colon cancer cell line models and xenografts [32]. The addition of TMZ to PARP inhibitors is believed to increase cell death not only by aggravating DNA damage, but also via increased PARP trapping [27, 33].

Topoisomerase I inhibitors (irinotecan and its active metabolite, SN-38) have also been combined with PARP inhibitors to demonstrate anti-tumour effects in preclinical models. Examples include with PARP inhibitors in *BRCA1/2* wildtype *in vitro* and xenograft models of triple negative breast cancer [34], with niraparib across *in vivo* and *in vitro* models of colorectal cancer without microsatellite instability [35], with veliparib in *in vitro* colon cancer models [36], with olaparib in colon cancer cell lines *in vitro* and in xenografts [37], and with PARP inhibitors in mouse leukemic cells [38].

Anti-tumour effects have also been elicited with combination of doxorubicin and olaparib in 2D and 3D ovarian cancer models [39], and between olaparib and a range of chemotherapies in oesophageal squamous cell carcinoma cell lines [40]. Most recently, paclitaxel was suggested to sensitise homologous recombination-proficient ovarian cancer cells to olaparib [41]. By contrast Murai and colleagues reported that PARP inhibitors have little combinatorial effect with cisplatin, but they synergise with camptothecin [33]. Studies of triplet treatment with chemotherapy, irradiation and PARP inhibition has also shown promise in colorectal cancer models [42].

This significant body of pre-clinical data has led to ∼80 clinical trials to test PARP inhibitor and chemotherapy combinations (see **Supplementary Table 1**) [43]. An assessment of these trials shows that combination PARP inhibitors and chemotherapy has primarily been assessed on unselected patient populations (**Figure 2**). In these trials only 22/79 (28%) included DNA repair function as an inclusion criterion for patients entering the trial. Thus, the majority of trials (72%; 57/79) do not require patients to have tumours with altered DNA repair status. 19/57 trials specify secondary analyses relating to DNA repair markers, however since these trials mostly occur across cancer types with low penetrance of DNA repair defects, these may not have accrued any cases with DNA repair alterations, or may not be sufficiently powered to draw any conclusions. Of the fourteen clinical trials that are actively recruiting or preparing to recruit patients, nine do not use DNA damage biomarkers as inclusion criteria, including trials on gastric and small cell lung cancer (NCT05411679), relapsed paediatric acute myeloid leukemia (NCT05101551), uterine leiomyosarcoma (NCT05432791), pancreatic cancer (NCT05257993), advanced stage rare cancers (NCT05142241), leiomyosarcoma and renal cell cancers (NCT04603365), breast cancer (NCT03150576), small cell cancers (NCT04209595), and ewing sarcoma and rhabdomyosarcoma (NCT01858168).

**Figure 2.**
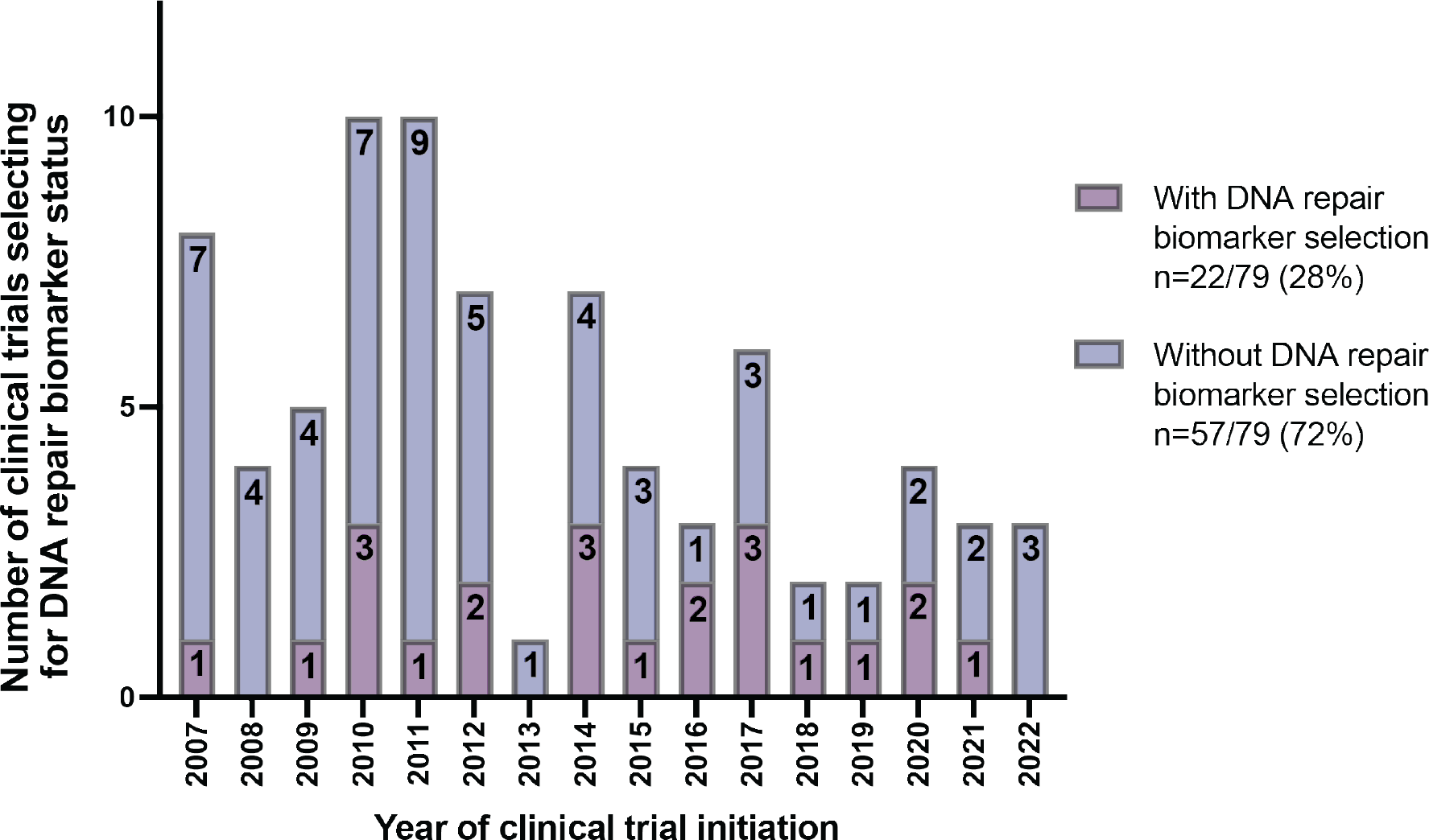
Number of combination PARP inhibitor and chemotherapy clinical trials selecting for DNA repair biomarker status and their year of initiation. 22/79 (28%) clinical trials included DNA repair biomarkers as a selection criterion for patients entering the trial and 57/79 (72%) clinical trials did not include DNA repair biomarkers as a selection criterion. Search of these clinical trials was completed in August 2022. Full details of trials are shown in Supplementary Table 1.

Outcomes of combination trials have now been reported, allowing an assessment of whether chemotherapy plus PARP inhibitor treatment generally improves outcome for patients. Importantly there are now sufficient studies to assess whether any improvement in survival is dependent on the presence of a DNA repair biomarker. Here, we provide the first synthesised analysis of the benefit of PARP inhibitors plus chemotherapies across multiple malignancies. Our study addresses whether the success of combination therapies in pre-clinical models without DNA repair vulnerabilities translates to an improvement in outcome of all patients.

## METHODS

### PRISMA statement

This meta-analysis was performed following the Preferred Reporting Items for Systematic reviews and Meta-analysis (PRISMA) statement.

### Search strategies

In October 2021, a systematic literature search of all English-language, phase II/III randomised controlled trials was conducted on PubMed, EMBASE and Conference Proceedings Citation Index – Science, with additional searches performed in August 2022. The website ClinicalTrials.gov was searched for additional clinical trials. The search terms used were “PARP-inhibitor”, “chemotherapy” and “combination”. To identify only randomised controlled trials during the search, we used the format of ‘Cochrane Highly Sensitive Search Strategy’ [44]. The reference lists of identified articles were also interrogated to further detect any relevant studies.

### Eligibility criteria

For progression-free survival (PFS), the inclusion criteria for the meta-analyses were: 1) phase II/III randomised controlled trials, 2) patients with cancer of any origin, 3) available biomarker data 4), patients were treated with PARP inhibitors in combination with chemotherapy in the experimental arm and chemotherapy plus placebo in the control arm, and 5) available information on PFS.

Exclusion criteria for PFS was studies where data could not be extracted on specified outcomes or DNA repair biomarkers.

For the meta-analyses of adverse effects, the inclusion criteria were the same as for PFS, except studies did not have to select for DNA repair biomarkers. Additional exclusion criteria for adverse effects were: 1) studies did not report adverse effects, and 2) studies did not report adverse effects as total per treatment arm and was therefore unable to be compared with other studies.

### Outcomes

The primary outcome was PFS defined as the time from randomisation to disease progression or death. Secondary endpoint was to determine common adverse effects and serious adverse effects (grade 3 and 4) of PARP inhibitors in combination with chemotherapy or chemotherapy alone.

### Data extraction

Based on inclusion and exclusion criteria, two reviewers independently identified studies and the following data was recorded: first author information, publication year, disease setting, study design, number of patients, DNA repair status of patients, type of intervention/control, therapeutic doses of treatment and control arms, and outcomes of interest including PFS, hazard ratios (HRs) and 95% confidence intervals (CIs), and the number of each of the selected adverse effects. In papers with multiple measures of DNA repair deficiency, we extracted data from subgroups that provided the greatest significant difference [15, 45, 46]. In the case where studies only provided Kaplan-Meier curves [47, 48], the software DigitizeIt was used to digitize the graph and extract the individual event and censoring times. The coxph function in R version 4.1.2 [49, 50] was used to calculate the HRs and their CIs from the reconstructed datasets. For the analyses of adverse effects, we recorded the number of patients with selected toxicities treated with PARP inhibitor and chemotherapy and total number of patients treated with PARP inhibitor and chemotherapy. We also extracted the number of patients with selected toxicities treated with chemotherapy alone and the total number of patients treated with chemotherapy alone.

### Risk-of-bias assessment

Two investigators independently assessed the quality of the studies included using version 2 of the Cochrane risk of bias tool for randomised trials (RoB 2) [51]. The risk of bias included bias arising due to the randomisation process, bias due to deviations from intended interventions, bias due to missing outcome data, bias in measurement of outcome and bias in selection of the reported result. Each category was judged as high, low or some concerns for risk of bias. Disagreements were resolved through discussion and consensus among the two reviewers.

### Statistical analysis

HR for PFS and 95% CI were extracted from each study or calculated based on survival curves of included papers. Pooled HRs were calculated by the generic inverse-variance method with a fixed-effects model, except as adjusted below following assessment of heterogeneity. Studies were weighted based on variance. For the analysis of risk of an adverse effect, patients with adverse effects assigned to the PARP inhibitor and chemotherapy arm were compared with those assigned to the chemotherapy alone control arm in the same trial for the calculation of risk difference.

Statistical heterogeneity was assessed using a Chi-squared (χ^2^) test and inconsistency index (I^2^) test. A fixed effects model was used when I^2^ value was less than 50%. Above 50%, a random effects model was used.

A forest plot was generated to visualise each study and pooled analysis. The weight of each study was represented as a square. CIs of each study were indicated as a line across the square. The pooled HR or risk difference was represented as a diamond. A p-value of < 0.05 was considered significant.

## RESULTS

### Study selection and characteristics for analysis of PFS

For examining the endpoint of PFS, 9 studies were included following the literature search. The PRISMA flow diagram summarises the process of identifying studies (**Figure 3)**. The flow chart presents number of studies identified, screened, included, and excluded, and the reasons for exclusion. Title and abstract screening identified 39 possible studies. Of these, 21 studies were excluded because of the following reasons: did not investigate PARP inhibitors combined with chemotherapies (immunotherapies, angiogenesis therapies), were not phase II/III randomised controlled trials (phase I/II safety trials), or only investigated DNA repair deficient (BRCA-mutated) cohorts. After full-text evaluation, 9 studies were excluded because of the following reasons: no data on DNA repair status of patients, unable to extract HR from data provided, data on PFS not available, or reporting of different endpoints of the same clinical trial across multiple publications (trials excluded [20, 52]) The risk-of-bias assessment of the included studies is presented in **Supplementary Figure 1**. Two studies raised concerns about the randomisation process and outcome measurements due to lack of information about the study, but no studies were excluded.

**Figure 3.**
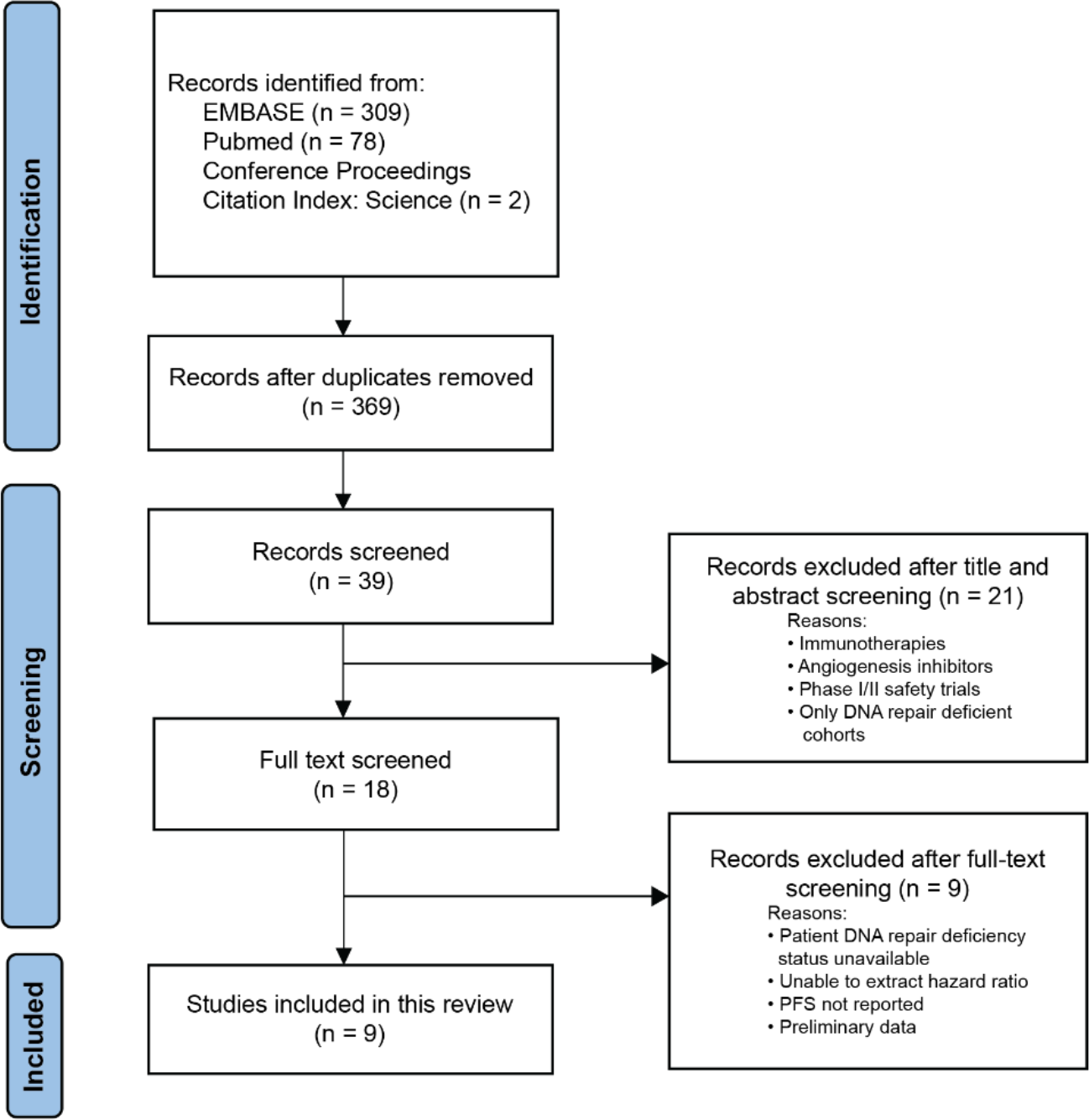
PRISMA flow diagram to identify included studies. PFS: progression-free survival.

The total sample size of the included studies for PFS analysis was 2,084 patients. All included studies reported the effect of PARP inhibitors in combination with chemotherapy versus chemotherapy alone in terms of PFS [15, 45–48, 53–56]. Three studies recruited patients with ovarian cancer, two with breast cancer, two with small cell lung cancer, one with gastric cancer and one with melanoma. The main characteristics of included studies are listed in **Table 2**.

**Table 2.** Characteristics of included studies.

| Study | Disease setting | Experimental arm | Control arm | DNA repair biomarker | DNA repair deficient | DNA repair competent | PARP inhibitor dose | Chemotherapy dose | Analyses |
| --- | --- | --- | --- | --- | --- | --- | --- | --- | --- |
| Coleman 2019 [15] | Metastatic high-grade serous ovarian cancer | Veliparib + Carboplatin/Paclitaxel | Carboplatin/Paclitaxel | *BRCA1/2* & HRD-score | 200 | 249 | Veliparib at a dose of 150 mg twice daily | Carboplatin given at AUC of 6 mg/ml/minute and paclitaxel 175 mg/m<sup>2</sup> | PFS and AE |
| Kummar 2015 [47] | Primary high-grade serous ovarian cancer | Veliparib + Cyclophosphamide | Cyclophosphamide | BRCAPRO score | 28 | 25 | Veliparib 60mg once daily | Cyclophosphamide 50mg once daily | PFS |
| Oza 2015 [56] | Primary high-grade serous ovarian cancer | Olaparib + Carboplatin/Paclitaxel | Carboplatin/Paclitaxel | *BRCA1/2* | 41 | 66 | Olaparib 200mg twice daily | Paclitaxel 175 mg/m<sup>2</sup> | PFS and AE |
| Geyer 2022 [54] | Primary triple-negative breast cancer | Veliparib + Carboplatin/Paclitaxel | Carboplatin/Paclitaxel | *BRCA1/2* | 70 | 406 | Veliparib 50mg twice daily | Carboplatin at AUC of 6 mg/ml/min and paclitaxel 80 mg/m<sup>2</sup> | PFS |
| Sharma 2020 [46] | Metastatic triple-negative breast cancer | Veliparib + Cisplatin | Cisplatin | *BRCA1/2* & HRD-score | 101 | 110 | Veliparib 300mg twice daily | Cisplatin 75mg/m<sup>2</sup> | PFS |
| Dieras 2020 [57] | Metastatic breast cancer | Veliparib + Carboplatin/Paclitaxel | Carboplatin/Paclitaxel | *BRCA1/2* | 509 | N/A | Veliparib 120mg twice daily | Carboplatin given at AUC of 6mg/mL/min and paclitaxel 80mg/m<sup>2</sup> | AE |
| Han 2020 [58] | Metastatic breast cancer | Veliparib + Carboplatin/Paclitaxel | Carboplatin/Paclitaxel | *BRCA1/2* | 294 | N/A | Veliparib 40mg twice daily | Carboplatin given at AUC 6mg/mL/min and paclitaxel 175mg/m<sup>2</sup> | AE |
| Pietanza 2018 [48] | Metastatic small cell lung cancer | Veliparib + Temozolomide | Temozolomide | SLFN11 | 25 | 23 | Veliparib 40mg twice daily | Temozolomide 200 mg/m<sup>2</sup> | PFS |
| Byers 2021 [53] | Metastatic small cell lung cancer | Veliparib + Carboplatin/Etoposide | Carboplatin/Etoposide | SLFN11 | 47 | 43 | Veliparib 240 mg twice daily | Carboplatin given at AUC of 5 mg/mL/min and etoposide 100 mg/m<sup>2</sup> | PFS and AE |
| Govindan 2022 [60] | Metastatic non-small cell lung cancer | Veliparib + Carboplatin/Paclitaxel | Carboplatin/Paclitaxel | N/A | 595 | N/A | Veliparib 120mg twice daily | Carboplatin given at AUC 6mg/mL/min and paclitaxel 200mg/m<sup>2</sup> | AE |
| Ramalingam 2021 [16] | Metastatic squamous non-small cell lung cancer | Veliparib + Carboplatin/Paclitaxel | Carboplatin/Paclitaxel | N/A | 970 | N/A | Veliparib 120mg twice daily | Carboplatin given at AUC 6mg/mL/min and paclitaxel 200mg/m<sup>2</sup> | AE |
| Liu 2018 [45], Bang 2017 [20] | Metastatic gastric cancer | Olaparib + Paclitaxel | Paclitaxel | ATM | 76 | 324 | Olaparib 100 mg twice daily | Paclitaxel 80 mg/m<sup>2</sup> | PFS and AE |
| Middleton 2015 [55] | Metastatic melanoma | Veliparib + Temozolomide | Temozolomide | ERCC1 | 129 | 121 | Veliparib 40 mg twice daily | Temozolomide 150 mg/m<sup>2</sup> | PFS |

HRD-score: homologous recombination deficiency score; PFS: progression-free survival; AE: adverse effect. Studies are organised based on cancer subtype.

Results were stratified for DNA repair status (deficient or competent). A patient was considered DNA repair deficient if they had an alteration (mutation, loss, or gain) of a gene involved in DNA repair that has been demonstrated to render increased sensitivity to PARP inhibitors. These included *BRCA1*, *BRCA2*, ATM, SFLN11 and ERCC1. Alternatively, a patient was deemed DNA repair deficient if they were greater than cut-off of 30% on the BRCAPRO test, or greater than 42 on ‘HRD-score” (Table 1).

A patient was deemed DNA repair competent if their cancer lacked alterations in the DNA repair marker in the study. Some studies used multiple markers of DNA repair status, and the marker used with respect to each study is specified in the following analyses. After these considerations, 717 patients were DNA repair deficient, and 1,367 patients were DNA repair competent for the purpose of the meta-analysis.

### Study selection and characteristics for analysis of adverse effects

For examining the endpoint of adverse effects, eight studies were included following the literature search. The PRISMA flow diagram summarises the process of identifying studies (**Figure 4)**. To identify these studies, we screened the literature using the same key words as for the primary meta-analysis but adjusted our inclusion/exclusion criteria to remove the requirement for DNA repair biomarkers. Title and abstract screening identified 39 possible studies. Of these, 21 studies were excluded because of the following reasons: did not investigate PARP inhibitors combined with chemotherapies (immunotherapies, angiogenesis therapies) or were not phase II/III randomised controlled trials (phase I/II safety trials). After full-text evaluation, seven studies were excluded because they did not provide data for adverse effects or were not comparable because they did not report patient totals per treatment arm. Note that four studies from our previous analyses on PFS also reported adverse effects that were able to be combined and compared, as in **Table 2**. Sharma et al [46] did not report adverse effects and the four other studies did not provide data for “any” adverse effects or “Any” grade 3/4 adverse effects. Since we were interested in tolerability of combination therapy regardless of underlying DNA repair function, we included an additional seven studies that reported adverse effects associated with combination PARP inhibition and chemotherapy that did not qualify for the meta-analysis on PFS. In total, we found eight studies that reported adverse effects on combination PARP inhibitor and chemotherapy compared to chemotherapy alone (**Table 2**). The risk-of-bias assessment of the included studies is presented in **Supplementary Figure 2**. Several studies were not double-blinded or were open label, leading to some concerns of bias, but were not excluded from the analysis.

**Figure 4.**
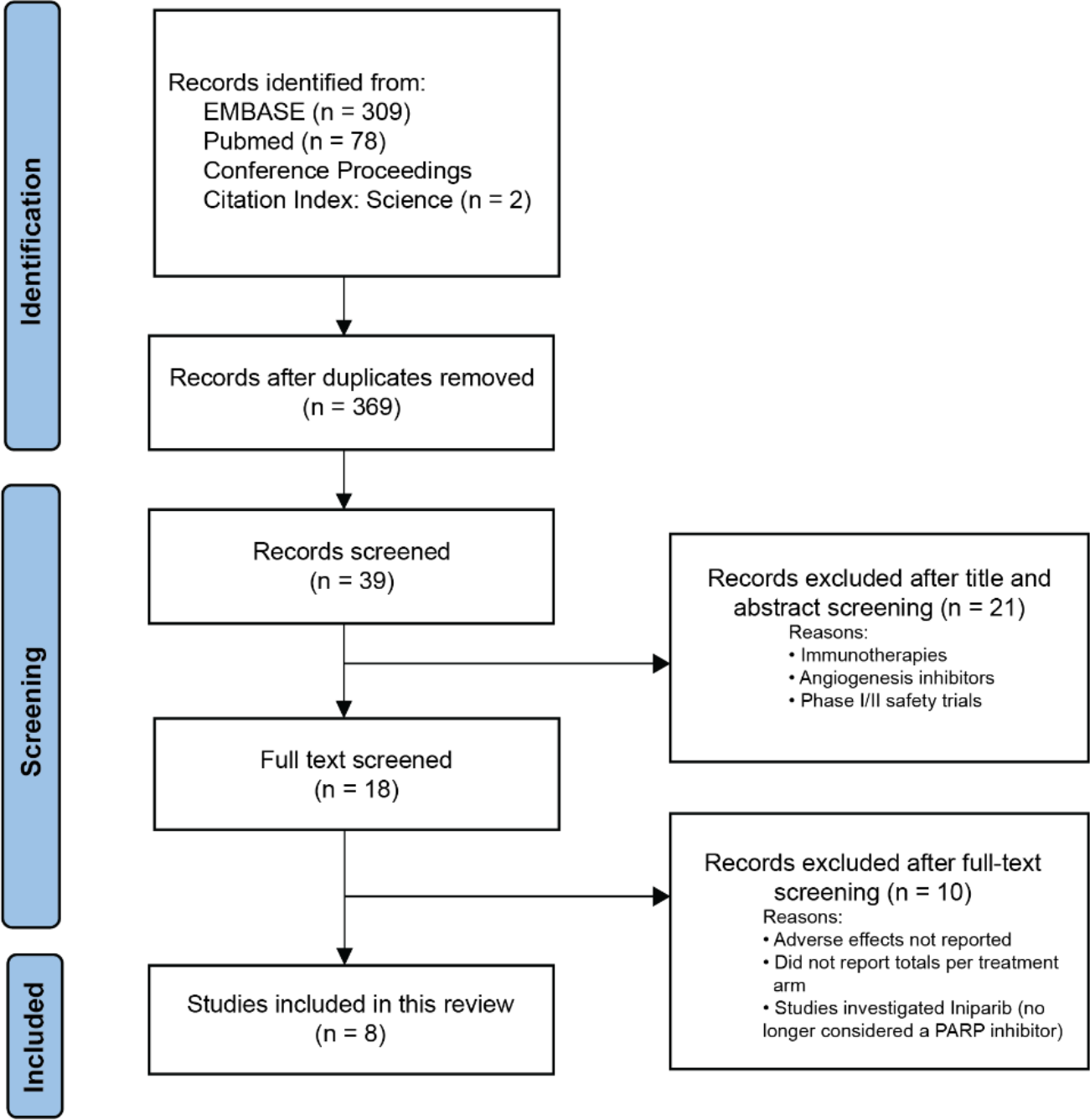
PRISMA flow diagram for identifying studies involving adverse effects of combination PARP inhibitor and chemotherapy.

The total sample size of the included studies for analysis of adverse effects was 4,498 patients. All included studies reported the adverse effects of PARP inhibitors in combination with chemotherapy vs chemotherapy alone, [15, 16, 45, 53, 56–62]. For analysis of adverse effects, a total of 2,347 patients were analysed in the combination PARP inhibitor and chemotherapy arm and a total of 2,151 patients in the chemotherapy alone arm. The main characteristics of included studies are listed in **Table 2**.

### DNA REPAIR DEFICIENT PATIENTS SHOW BENEFIT IN PFS, BUT DNA REPAIR COMPETENT PATIENTS DO NOT

To evaluate the effect of PARP inhibitor in combination with chemotherapy in DNA repair deficient patients, we conducted a pooled analysis to compare the PFS between PARP inhibitor in combination with chemotherapy and the chemotherapy alone group. In total, all 9 studies were pooled for this analysis. For our analysis, a patient was considered DNA repair deficient if they had an alteration of a gene involved in DNA repair that has been demonstrated to render increased sensitivity to PARP inhibitors (Table 1). For Byers (2021) and Pietanza (2018), we considered SLFN11-positive patients as DNA repair deficient, and SLFN11-negative patients as DNA repair competent [52]. For Geyer (2022), Oza (2015) and Kummar (2015), patients with a *BRCA1/2* mutation were considered DNA repair deficient and those with no mutation in *BRCA1/2* were considered DNA repair competent [53,55]. Additionally, in Kummar (2015), patients were considered DNA repair deficient if they had a BRCAPRO score of >30%. For Liu (2018), ATM-negative patients were considered DNA repair deficient and ATM-positive patients were considered DNA repair competent [44]. For Middleton (2015), low ERCC1 expression was considered DNA repair deficient and high ERCC1 expression was considered DNA repair competent [54].

In the Coleman (2019) and Sharma (2020) studies, there were multiple biomarkers of impaired DNA repair. For analysis of Coleman (2019), we used “BRCA-mutated” (patients who had germline or tissue-based mutations as determined by the Myriad BRACAnalysis CDx and myChoice homologous recombination deficiency CDx assay, respectively, in *BRCA1* or *BRCA2)* cohort as our “DNA repair deficient” and the “non-homologous recombination deficiency” (no genetic evidence of homologous recombination deficiency) cohort as “DNA repair competent” [15]. For Sharma (2020), patients were classified into “BRCA-like” (included myChoice homologous recombination deficiency score, somatic *BRCA1/2* mutations, BRCA1 methylation and non-*BRCA1/2* homologous recombination germline mutations) and “non-BRCA-like”. For our analysis, we defined “BRCA-like” as DNA repair deficient, and “non-BRCA-like” as DNA repair competent.

717 DNA repair deficient patients received either PARP inhibitor in combination with chemotherapy or chemotherapy alone. Pooled analysis on the DNA repair deficient population showed that PARP inhibitor in combination with chemotherapy, compared to chemotherapy alone, was significantly associated with improved PFS (HR:0.52, 95% CI: 0.43-0.63, p < 0.00001) (**Figure 5A**).

**Figure 5.**
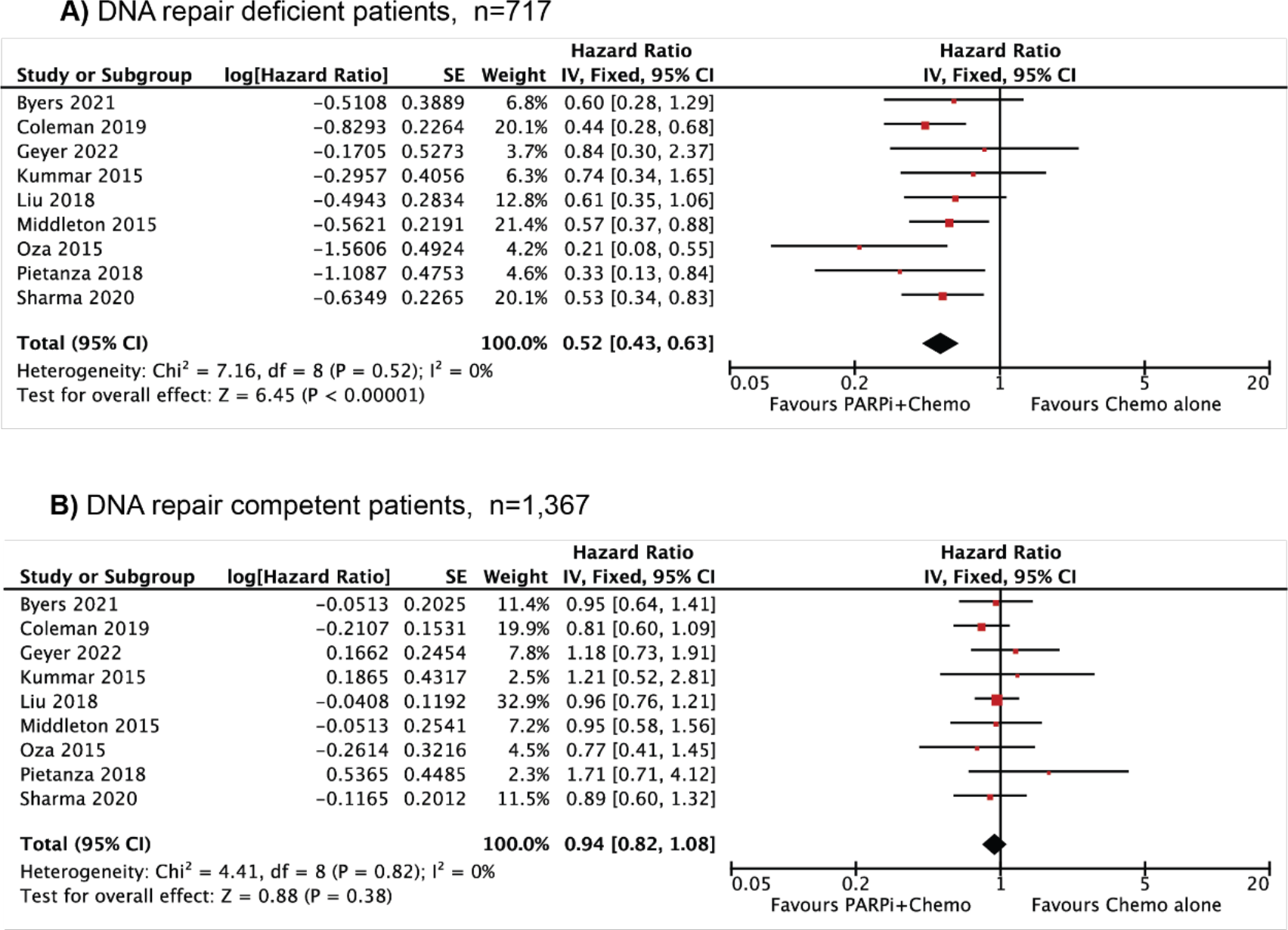
Comparative effects of combination PARP inhibitor and chemotherapy vs chemotherapy alone in A) DNA repair deficient patients (n=717) and in B) DNA repair competent patients (n=1,367). Results were presented as individual and pooled hazard ratio (HR) with 95% confidence interval (CI). SE: standard error; IV: inverse-variance.

1,367 DNA repair competent patients were pooled for analysis. No significant difference in PFS was observed in DNA repair competent patients with PARP inhibitor in combination with chemotherapy compared to chemotherapy alone (HR: 0.94, 95% CI: 0.81-1.08, p = 0.38) (**Figure 5B**).

There was some heterogeneity in these studies including DNA repair deficient patients (Chi^2^=7.16), although this was non-significant, and the inconsistency score (I^2^) was 0%.

In the subsequent sections we performed subgroup analyses to determine if there were any factors, including cancer subtype, DNA repair biomarker, or therapy subtype, which could be significantly contributing to the heterogeneity of response.

## 1. DNA REPAIR STATUS DICTATES BENEFIT, REGARDLESS OF CANCER SUBGROUP

We next analysed response within cancer subtypes to determine if benefit from combination therapy could be mainly attributed to certain cancer subtypes. We pooled studies that had two or more trials investigating the same cancer type. The combination therapy significantly improved PFS in DNA repair deficient patients in ovarian cancer (HR: 0.44, 95% CI: 0.31- 0.64, p < 0.00001), breast cancer (HR: 0.57, 95% CI: 0.38-0.86, p = 0.007) and small cell lung cancer (HR: 0.47, 95% CI: 0.26-0.85, p = 0.01) (**Figure 6A(i), B(i), C(i)**). In contrast, DNA repair competent patients had no associated improvement in PFS from the combination therapy in any of the cancer subtypes (**Figure 6A(ii), B(ii), C(ii)**).

**Figure 6.**
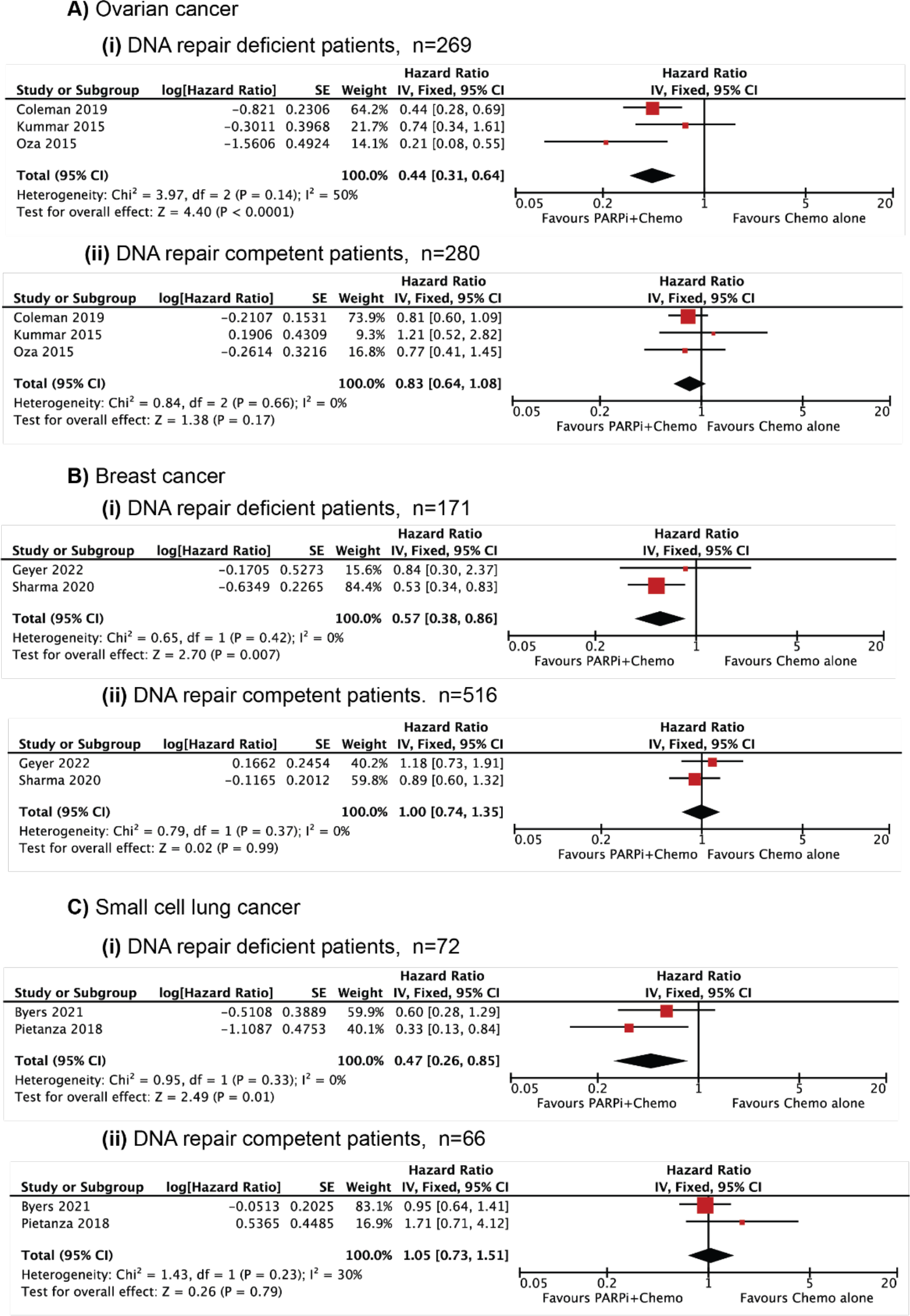
Comparative effects of combination PARP inhibitor and chemotherapy vs chemotherapy alone separated by cancer subgroup. A) Ovarian cancers with (i) DNA repair deficient patients (n=269) and (ii) DNA repair competent patients (n=280). B) Breast cancers with (i) DNA repair deficient patients (n=171) (ii) DNA repair competent patients (n=516). C) Small cell lung cancers with (i) DNA repair deficient patients (n=72) and (ii) DNA repair competent patients (n=66). Results were presented as individual and pooled hazard ratio (HR) with 95% confidence interval (CI). SE: standard error; IV: inverse-variance.

## 2. DNA REPAIR FUNCTION STATUS DICTATES BENEFIT, REGARDLESS OF BIOMARKER SUBGROUP

We next looked at pooled biomarkers to explore whether benefit from PARP inhibitor in combination with chemotherapy was only associated with certain biomarkers of DNA repair function. First, we examined studies that incorporated patients with “*BRCA*-like” biology. These studies included patients that were categorised based on *BRCA1/2* mutation or a surrogate marker to categorise cancers as DNA repair deficient due to a ‘*BRCA*-like’ event. Two papers in this analysis incorporated surrogate *BRCA*-like markers [15, 46]. Sharma (2020) defined the ‘*BRCA*-like’ group with 4 markers: homologous recombination deficiency genomic instability score of > 42, somatic *BRCA1/2* mutation, *BRCA1* promoter methylation, and germline DNA repair gene mutations other than *BRCA1/2*. Positivity on any 1 of the 4 markers placed a patient in the *BRCA*-like group. ‘Non-*BRCA-*like’ patients included patients that did not have any of the 4 markers above. In Coleman (2019), multiple measures of *BRCA1/2* status were compared including germline *BRCA1/2* mutation (gBRCAmut), and ‘*BRCA*-like’ patients include those that have somatic and germline *BRCA1/2* gene mutations and/or a ‘*BRCA*-like’ phenotype based on DNA repair deficiency assays. We extracted data from ‘gBRCAmut’ for the purpose of this subgroup analysis, as Coleman *et al* concluded that it was a better discriminator for improvement in survival based on their analyses.

*BRCA*-like patients showed significant improvement in PFS with the combination therapy (HR: 0.50, 95% CI: 0.38-0.65, p < 0.00001) (**Figure 7A**). There was no significant difference in the PFS of non-BRCA-like patients with the addition of PARP inhibitor (HR: 0.90, 95% CI: 0.74- 1.09, p = 0.29) (**Figure 7B)**. There was some heterogeneity observed in the analysis of the *BRCA-*like, DNA repair deficient patients (Chi^2^ = 5.43, I^2^ = 26%) likely due to the variability of definition of “*BRCA*-like” in these studies, although this was not enough to perform a random effects model.

**Figure 7.**
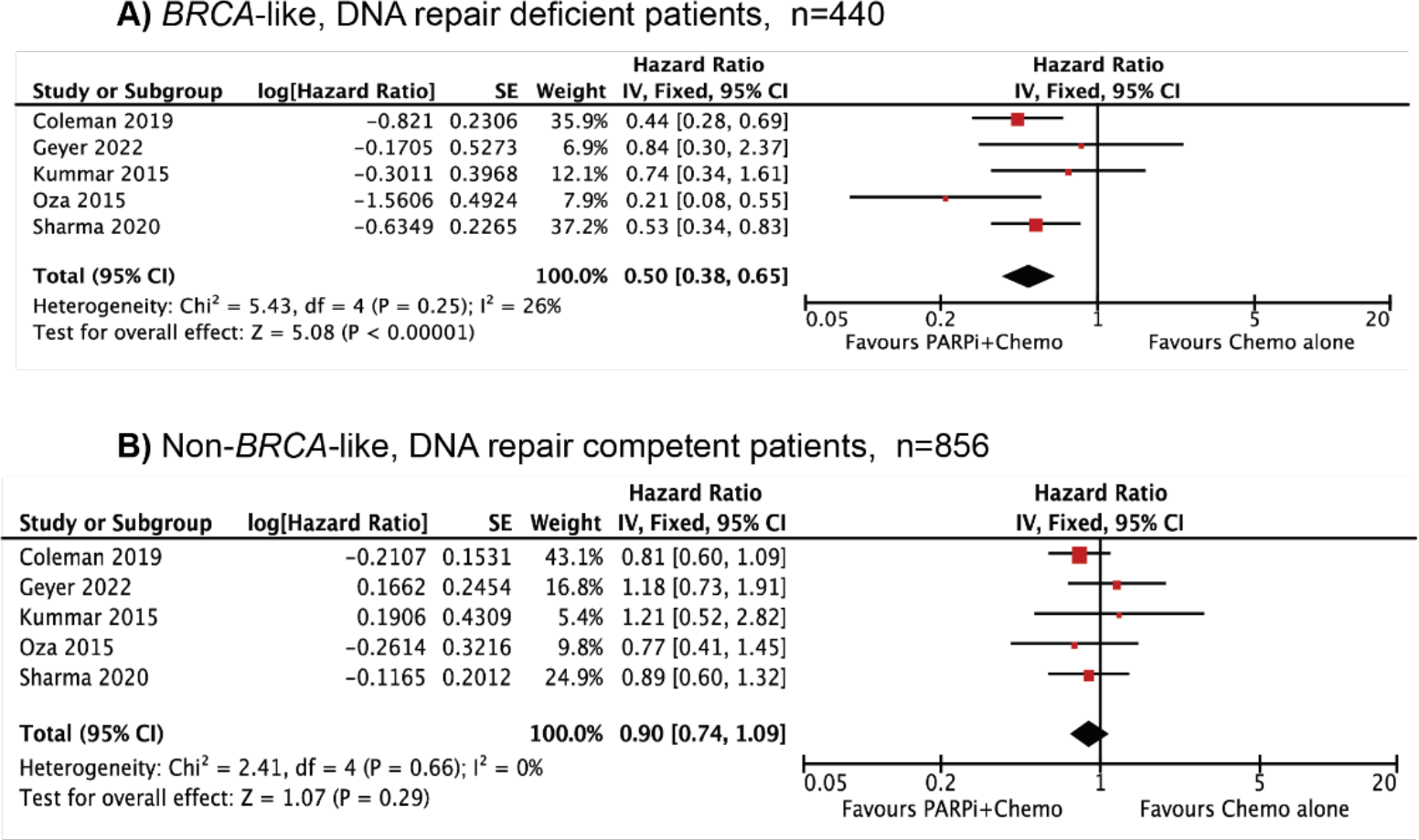
Comparative effects of combination PARP inhibitor and chemotherapy vs chemotherapy alone separated by *BRCA* status. A) DNA repair deficient, *BRCA*-like patients (n=440) B) DNA repair competent, non-*BRCA-*like patients (n=856). Results were presented as individual and pooled hazard ratio (HR) with 95% confidence interval (CI). SE: standard error; IV: inverse-variance.

Both studies of small cell lung cancer were performed with the biomarker SFLN11. High expression of SFLN11 was associated with increased PFS in patients treated with combination therapy (**Figure 6C(i), (ii)**). Patients with absent SFLN11 did not show any improvement in PFS with combination therapy.

Overall, the *BRCA*-like and SFLN11 biomarkers were both able to discriminate patient populations with an improved PFS when treated with combination therapy.

## 3. INVESTIGATION OF THE BENEFIT ASSOCIATED WITH SPECIFIC PARP INHIBITORS OR CHEMOTHERAPIES USED IN COMBINATIONS

We next considered whether the type of PARP inhibitor or the type of chemotherapy could influence the benefit seen in PFS following combination therapy. Two PARP inhibitors were used in these studies, veliparib (7 studies) [15, 46–48, 53–55] and olaparib (2 studies) [45, 56]. Notably, there was variability in dosing, scheduling, and length of treatments. Veliparib and olaparib were given twice daily, except for one study where veliparib was given once daily [47]. Doses of veliparib ranged from 20mg to 400mg and olaparib ranged from 100mg to 400mg. The length of treatment cycles varied between studies from 14-days to 28-day cycles. The number of cycles patients received also varied from patients receiving 1 cycle of treatment, to patients receiving 30 cycles of treatment.

A significant benefit to PFS was seen for DNA repair deficient patients treated with combination therapy regardless of the type of PARP inhibitor, whereas no benefit was seen in DNA repair competent patients (**Figure 8)**. Even though the conditions of treatment were highly variable, there was little heterogeneity in studies that included veliparib treatment (DNA repair deficient: Chi^2^=3.42, DNA repair competent: Chi^2^=3.99), and the inconsistency score (I^2^) was 0%. Only two studies examined olaparib treatment which probably led to the considerable heterogeneity (I^2^ =72%) between those studies as they were conducted on different cancer types with different treatment regimes. Consequently, we applied a random effects model to this analysis, and to the other analyses of olaparib for consistency.

**Figure 8.**
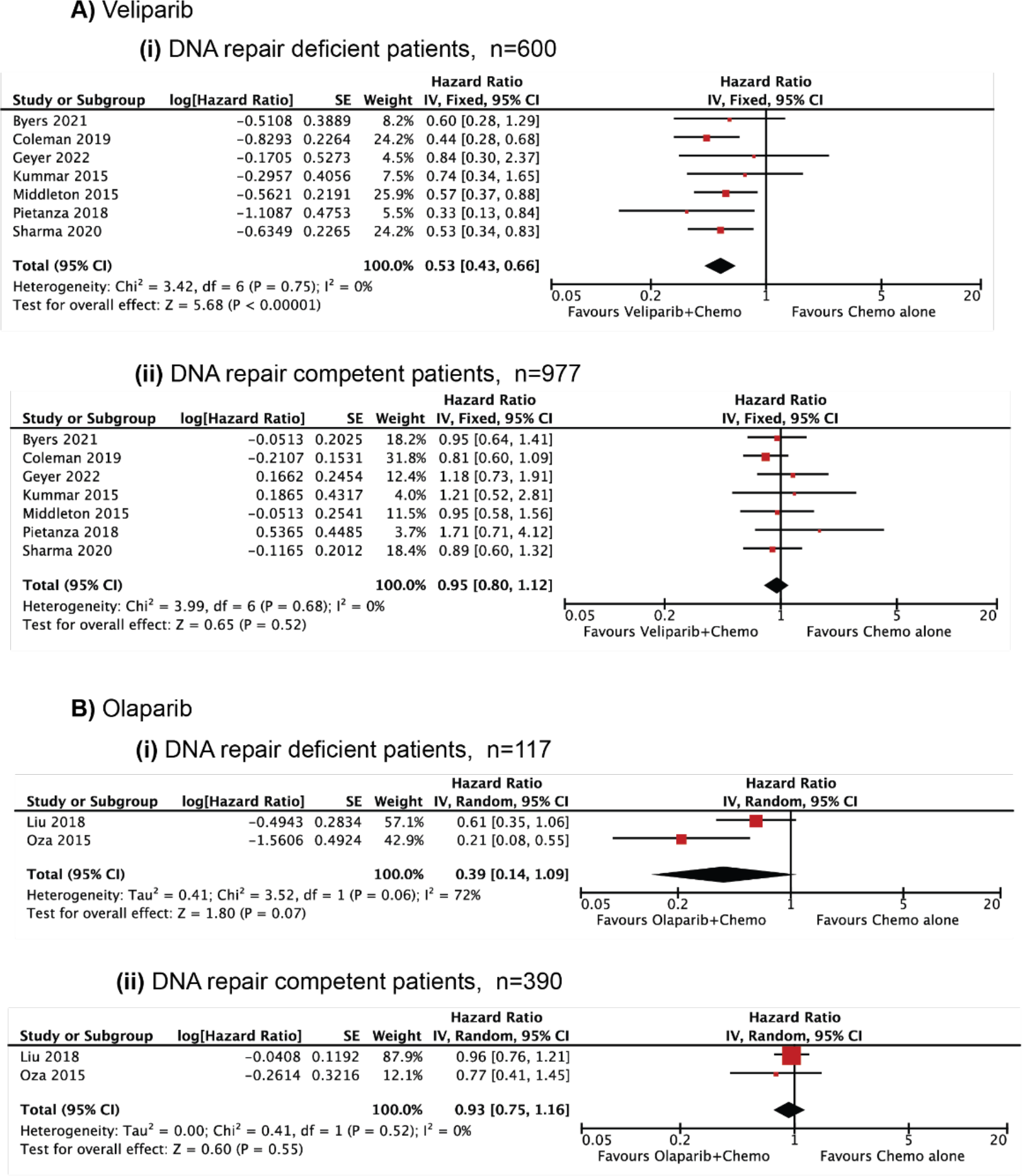
Comparative effects of combination PARP inhibitor and chemotherapy vs chemotherapy alone separated by type of PARP inhibitor. A) Veliparib in DNA repair deficient patients (n=600) B) Veliparib in DNA repair competent patients (n=977) C) Olaparib in DNA repair deficient patients (n=117) D) Olaparib in DNA repair competent patients (n=390). A) and B) were analysed using fixed effects model. C) and D) were analysed using random effects model. Results were presented as individual and pooled hazard ratio (HR) with 95% confidence interval (CI). SE: standard error; IV: inverse- variance.

The chemotherapy treatment across the 9 trials was highly varied, and included carboplatin, etoposide, paclitaxel, cyclophosphamide, and etoposide. Each trial also used different doses and schedules of chemotherapy treatment. However, the trials could be broadly separated into trials that included platinum-based therapies (carboplatin and cisplatin) and non-platinum therapies (cyclophosphamide, temozolomide and paclitaxel). As DNA repair deficient cancers are frequently more sensitive to platinum-based drugs [63], we considered whether the benefit of combination therapy is altered with this treatment.

A significant benefit to PFS was seen for DNA repair deficient patients treated with either platinum-based or other combination therapies. There was no improvement to PFS in DNA repair competent patients regardless of whether the chemotherapy included platinum-based drugs (**Figure 9**).

**Figure 9.**
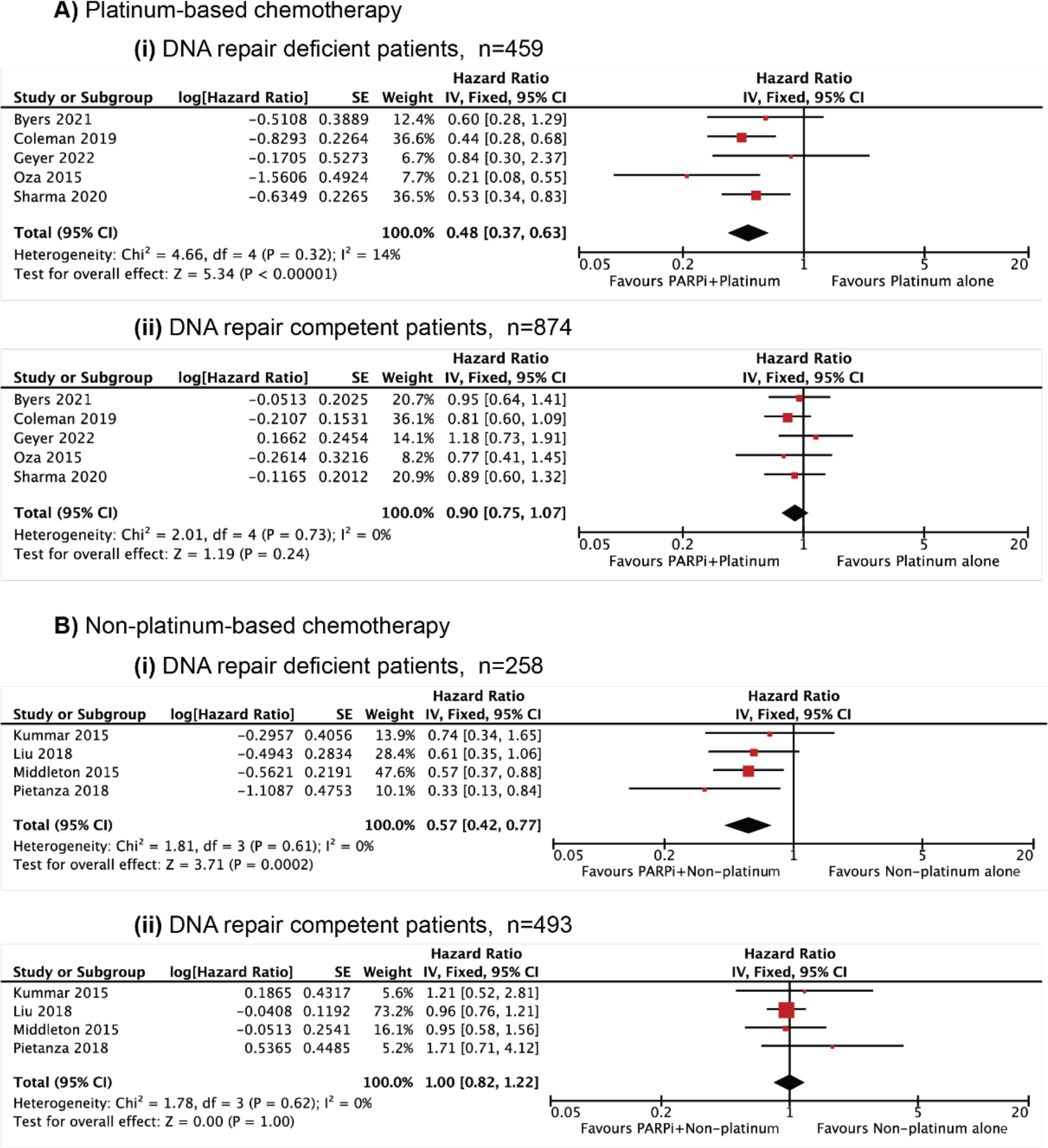
Comparative effects of combination PARP inhibitor and chemotherapy vs chemotherapy alone separated by type of chemotherapy. A) Platinum-based chemotherapy in (i) DNA repair deficient patients (n=459) and (ii) DNA repair competent patients (n=874). B) Non-platinum-based chemotherapy in (i) DNA repair deficient patients (n=258) and (ii) DNA repair competent patients (n=493). Results were presented as individual and pooled hazard ratio (HR) with 95% confidence interval (CI). SE: standard error; IV: inverse-variance.

## 4. ADVERSE EFFECTS: NO OVERALL INCREASE IN THE FREQUENCY OF ADVERSE EFFECTS, BUT SIGNIFICANTLY HIGHER GRADE 3/4 EFFECTS IN COMBINATION TREATED PATIENTS

Next, we assessed the secondary endpoint of whether the combination of PARP inhibitors and chemotherapy is well tolerated compared to chemotherapy alone. We found eight studies that reported adverse effects on combination PARP inhibitor and chemotherapy compared to chemotherapy alone. In total, we assessed 3.789 patients with reported adverse effects. 1,987 patients were treated with combination PARP inhibitor and chemotherapy and 1,802 patients were treated with chemotherapy alone (**Table 3**).

**Table 3.**
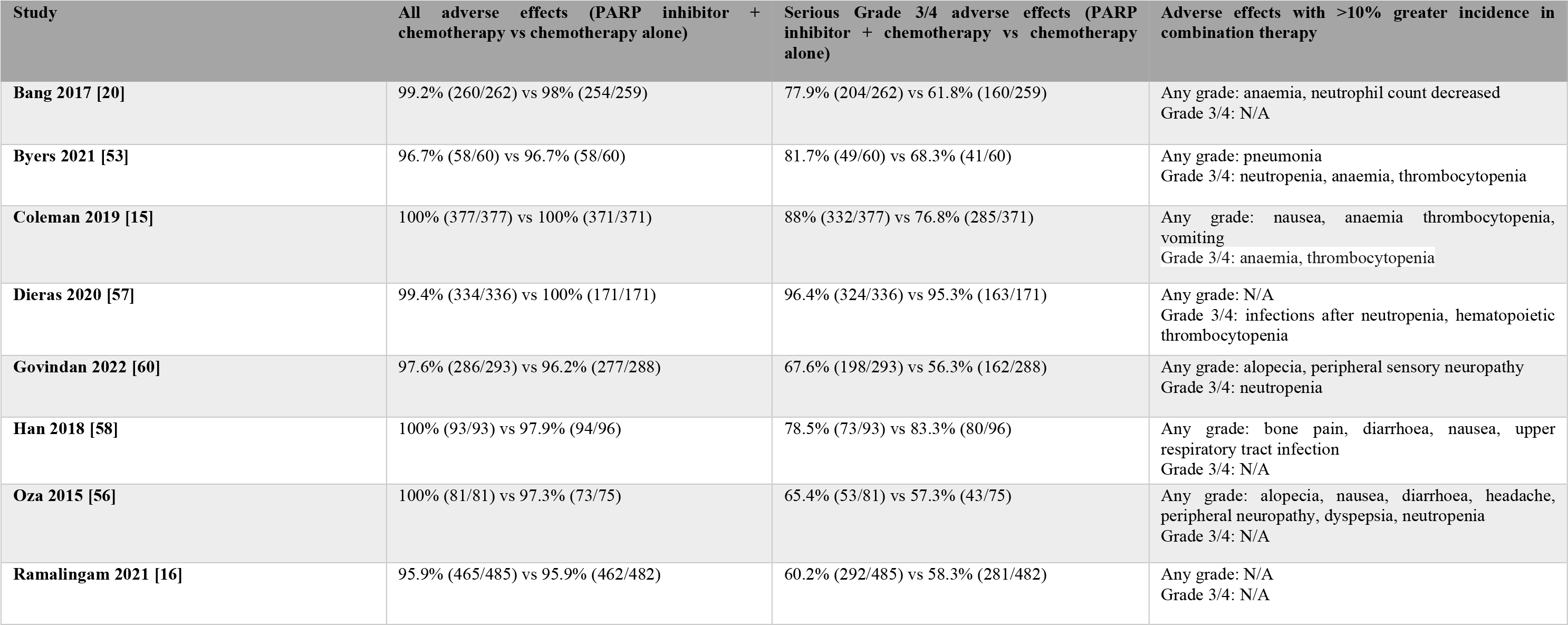
Characteristics of studies included in adverse effects analysis. N/A: not available.

| Study | All adverse effects (PARP inhibitor + chemotherapy vs chemotherapy alone) | Serious Grade 3/4 adverse effects (PARP inhibitor + chemotherapy vs chemotherapy alone) | Adverse effects with >10% greater incidence in combination therapy |
| --- | --- | --- | --- |
| **Bang 2017 [20]** | 99.2% (260/262) vs 98% (254/259) | 77.9% (204/262) vs 61.8% (160/259) | Any grade: anaemia, neutrophil count decreased<br>Grade 3/4: N/A |
| **Byers 2021 [53]** | 96.7% (58/60) vs 96.7% (58/60) | 81.7% (49/60) vs 68.3% (41/60) | Any grade: pneumonia<br>Grade 3/4: neutropenia, anaemia, thrombocytopenia |
| **Coleman 2019 [15]** | 100% (377/377) vs 100% (371/371) | 88% (332/377) vs 76.8% (285/371) | Any grade: nausea, anaemia thrombocytopenia, vomiting<br>Grade 3/4: anaemia, thrombocytopenia |
| **Dieras 2020 [57]** | 99.4% (334/336) vs 100% (171/171) | 96.4% (324/336) vs 95.3% (163/171) | Any grade: N/A<br>Grade 3/4: infections after neutropenia, hematopoietic thrombocytopenia |
| **Govindan 2022 [60]** | 97.6% (286/293) vs 96.2% (277/288) | 67.6% (198/293) vs 56.3% (162/288) | Any grade: alopecia, peripheral sensory neuropathy<br>Grade 3/4: neutropenia |
| **Han 2018 [58]** | 100% (93/93) vs 97.9% (94/96) | 78.5% (73/93) vs 83.3% (80/96) | Any grade: bone pain, diarrhoea, nausea, upper respiratory tract infection<br>Grade 3/4: N/A |
| **Oza 2015 [56]** | 100% (81/81) vs 97.3% (73/75) | 65.4% (53/81) vs 57.3% (43/75) | Any grade: alopecia, nausea, diarrhoea, headache, peripheral neuropathy, dyspepsia, neutropenia<br>Grade 3/4: N/A |
| **Ramalingam 2021 [16]** | 95.9% (465/485) vs 95.9% (462/482) | 60.2% (292/485) vs 58.3% (281/482) | Any grade: N/A<br>Grade 3/4: N/A |

Pooled data were stratified for all adverse effects and data was analysed using risk difference. We note that there are inconsistencies in the reporting of adverse effects (reported variously as occurring in >5% vs >10% vs >20% of patients), and different adverse effects were reported in each study. This is likely to have contributed to heterogeneity within the analyses. Our initial analysis demonstrated high heterogeneity in the analysis of grade 3/4 adverse effects (I^2^ = 74%). Consequently, we applied a random effects model to this analysis, and to the other analyses of adverse effects for consistency.

There was no significant difference in the incidence of all adverse effects between patients treated with combination therapy or chemotherapy alone (RD: 0.00, 95% CI: -0.01-0.01, p = 0.47) (**Figure 10A**). However, when looking at more severe side effects, the combination therapy was associated with a significant increase in the incidence of grade 3/4 adverse effects (RD: 0.07, 95% CI: 0.02-0.12, p = 0.009) (**Figure 10B)**. The high heterogeneity observed with the assessment of grade 3/4 effects led us to perform a more refined analysis on haematological effects, as these were very common and reported with greater consistency among the studies. We chose to investigate neutropenia as the most reported haematological effect. We found that neutropenia had increased incidence with the combination therapy (RD: 0.06, 95% CI: 0.03- 0.09, p = 0.0004) (**Figure 10C**). The analysis of neutropenia alone did not exhibit high heterogeneity.

**Figure 10.**
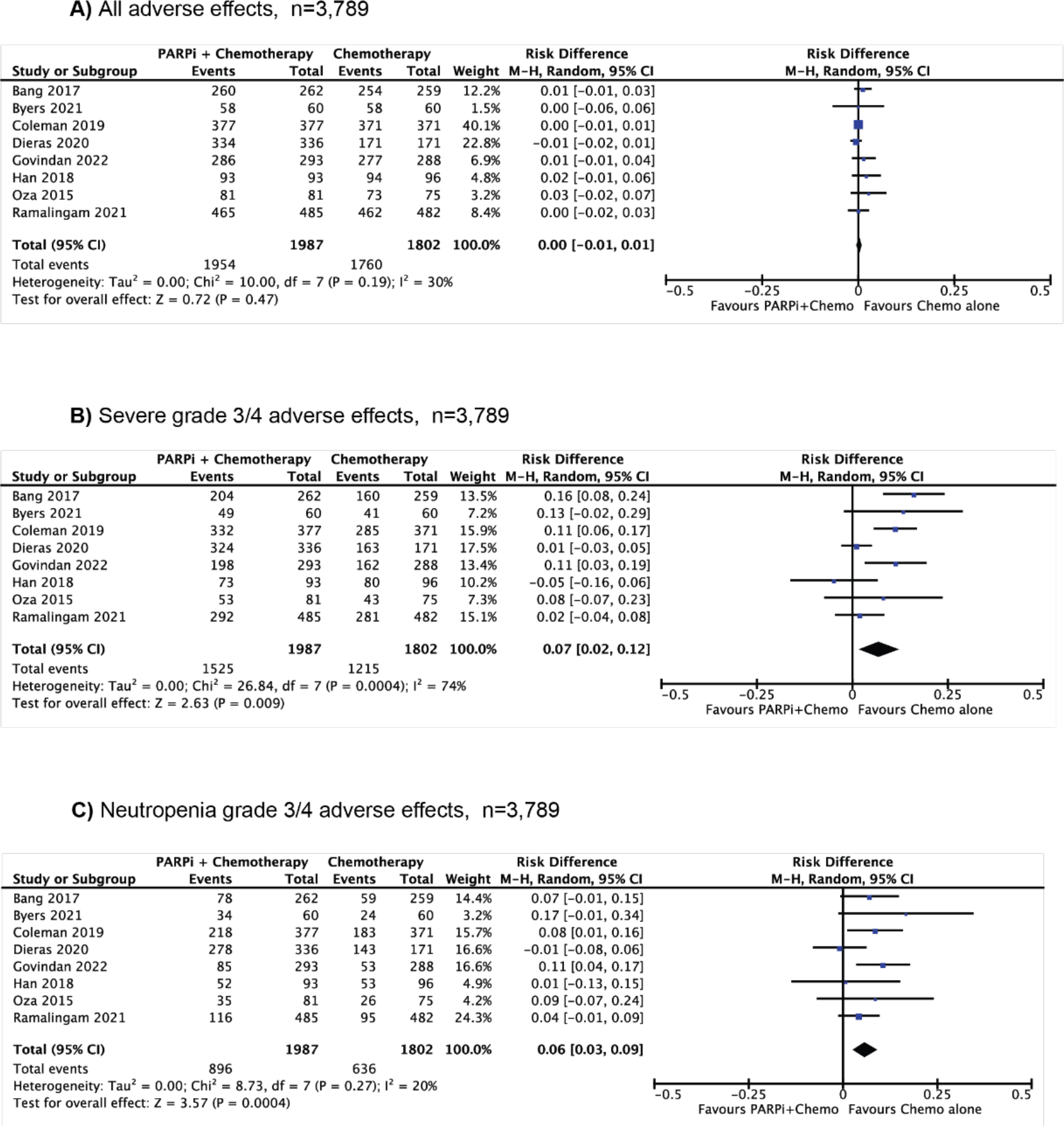
Adverse effects of combination of PARP inhibitor and chemotherapy vs chemotherapy alone. A) All adverse effects B) Severe grade 3/4 adverse effects C) Neutropenia grade 3/4 adverse effects (n=3,789). Results were presented as individual and pooled risk difference with 95% confidence interval (CI). M-H: Mantel-Haenszel.

## DISCUSSION

Several PARP inhibitors are approved by Food and Drug Administration (FDA) and European Medicines Agency (EMA) as maintenance therapies following first-line treatments for DNA repair deficient ovarian, breast, pancreatic and prostate cancers. PARP inhibitors are yet to be approved for use as part of short-term therapeutic regimens with curative intent in any malignancy. However, this is an intense area of clinical investigation with a multitude of clinical trials assessing the potential of combining PARP inhibitors with immunotherapies, radiation therapy, chemotherapies, and other therapies to improve rates of success of those therapeutic regimes.

Our results, combining nine studies and 2,084 patients across several cancer subtypes, demonstrates that patients with an identifiable DNA repair deficiency may gain an improvement in PFS if PARP inhibitors are combined with chemotherapy as a line of therapy. In contrast, DNA repair competent patients did not show any additional benefit (**Figure 5**). Furthermore, we found that benefit for DNA repair deficient patients was associated with combination therapy in all cancer subtypes investigated including ovarian, breast and small cell lung cancers (**Figure 6**). We considered whether certain biomarkers (*BRCA*-like, SLFN11) would have differences in their associated benefit, but found similar benefit was seen with the combination therapy in DNA repair deficient but not DNA repair competent patients regardless of the biomarker (**Figure 7**). We observed that the benefit for DNA repair deficient patients was not influenced by either the type of PARP inhibitor (veliparib, olaparib) or chemotherapy (platinum-based, non-platinum-based) (**Figure 8&9**).

All subgroup analyses supported the same conclusion that improved PFS from combination therapy was only associated with DNA repair deficiency status. However, there were several limitations to our study. First, each study involved different chemotherapies, PARP inhibitor and dosing regimens. This likely contributed to the heterogeneity observed across some of the subgroup analyses and analysis of adverse effects. To address issues of heterogeneity, we applied a random effects model to analyses where heterogeneity was greater than 50%, but the addition of more studies would be informative. A second limitation of this meta-analysis is that we were unable to assess the impact of treatment and DNA repair deficiency status on overall survival, as those data are not yet available due to the immaturity of trials in this area.

While this meta-analysis did not show any benefit in DNA repair competent cancers, it should also be considered whether the most optimal PARP inhibitors and chemotherapies were used in the combination therapies in these trials. Of note, both of the PARP inhibitors used in these analysed studies, veliparib and olaparib, have weak PARP trapping ability [64, 65]. Synergy with chemotherapy has been suggested to be dependent upon PARP trapping [27], and the combination of chemotherapy with efficient PARP trappers such as talazoparib [66] may provide greater synergistic effects. A drawback of PARP inhibitors with higher trapping ability, is the potential for greater challenges with toxicity. However, a recent phase I study investigated the safety of combined talazoparib and carboplatin/paclitaxel and found that the combination is feasible with an intermittent, lower dose schedule of talazoparib and appropriate supportive care [67]. Further investigation into the dosing regimen may assist in enhancing anti-tumour efficacy whilst improving tolerability.

PARP inhibitors are only approved for DNA repair deficient cancers. The exception is niraparib, which is approved as first line maintenance therapy regardless of homologous recombination deficiency status for ovarian cancers that have shown a response to platinum therapies. Response to platinum salts is correlated with the presence of homologous recombination deficiency [68], and has high penetrance in ovarian cancer with up to 69% classified as homologous recombination deficient [69]. Thus, it may be that the high penetrance of homologous recombination deficiency in ovarian cancers leads to a more general susceptibility to PARP inhibitors, alone or in combination with chemotherapies. Interestingly, from our subgroup analysis of ovarian cancers without a homologous recombination deficiency, there was an overall weighted average effect of 1.38 with combination therapy, although this was not significant (p<0.17; HR: 0.83; CI 0.64-1.08). Since this subgroup analysis only includes 340 patients over three studies, the addition of further studies may indicate benefit for combination PARP inhibitors and chemotherapy in ovarian cancer patients that do not have canonical homologous recombination deficiency indicators such as *BRCA1*/*BRCA2* mutation. This should be re-examined if more clinical trials are reported on PARP inhibitor/chemotherapy combinations for ovarian cancer.

A major consideration in this analysis is that the definition of DNA repair deficiency is highly heterogeneous, with individual clinical trials using different methods and cut-offs to assess a patient’s DNA repair deficiency status. This raises the question of whether there should be a uniform definition of DNA repair deficiency, or if each cancer subtype will require its own specific biomarkers. SFLN11 is an emerging biomarker of PARP inhibitor sensitivity, but unlike mutations to homologous recombination pathway genes it does not appear to be associated with the presence of genomic scars [22]. Instead SFLN11 is actively recruited to sites of DNA damage to inhibit homologous recombination [70], and its high expression is correlated with PARP inhibitor sensitivity [22]. This seems to be particularly relevant for small cell lung cancer [53], but a retrospective analysis of 110 ovarian cancers treated with PARP inhibitor suggests there could be broader application [71]. In that study, *BRCA*-wildtype ovarian cancer patients with high SFLN11 showed non-significant improvements in olaparib response compared to SFLN11 low patients [71]. Overall, each of these biomarkers should be assessed in each cancer setting for its frequency, overlap with other DNA repair biomarkers, and its potential as an indicator of PARP inhibitor sensitivity.

An area that was not explored with this meta-analysis was whether non-DNA repair markers may be predictive of benefit from combination PARP inhibitors and chemotherapy. In the study of Ramalingam *et al* [16] the LP52, a 52 gene panel that was first generated to differentiate squamous non-small cell lung cancer subtypes, was predictive of a better response to veliparib in combination with carboplatin/paclitaxel. A similar, but non-significant, trend was seen in the study of Govindan *et al* on veliparib with carboplatin/paclitaxel versus physician’s choice of standard chemotherapy in advanced non-squamous non-small cell lung cancer [60]. Thus, alternative markers may provide more guidance for combination therapy, especially in those cancer types without a high penetrance of DNA repair deficiencies.

Our secondary analysis of the adverse effects associated with combination treatment versus chemotherapy alone revealed that combination therapy had higher associated toxicities. Grade 3/4 adverse effects and neutropenia were significantly higher with chemotherapy and PARP inhibitor combination therapy, although there was no increase in overall effects of all grades. A confounding issue was the significant heterogeneity we observed when considering grade 3/4 adverse effects (**Figure 10B**). This was most likely due to the differences in criteria used for adverse effects in each clinical trial. Re-analysis of the data for neutropenia alone demonstrated an increased risk with combination therapy, and the data showed very little heterogeneity for this sub-analysis. Data from further clinical trials are needed to resolve whether other haematological and non-haematological side-effects are significantly associated with the combination of chemotherapy and PARP inhibitors. Despite these side-effects, it is important to note that the PFS benefit with the combination of PARP inhibitor and chemotherapy was achieved despite dose modifications occurring in most of the studies. Most studies found that the combination therapy had a manageable toxicity profile.

In conclusion, DNA repair deficiency associates with improved PFS following combination PARP inhibitor and chemotherapy compared to chemotherapy alone, but there is no associated benefit for DNA repair competent patients. Despite the potential to improve outcome for DNA repair deficient patients, the benefit must be weighed against the increase in associated toxicities. Results show that combined PARP inhibitor and chemotherapy are associated with an increase in severe adverse effects, particularly neutropenia, compared to chemotherapy alone. Previous meta-analyses have shown that haematological side-effects are very common with PARP inhibitors [72], and this effect is likely to be compounded with the addition of chemotherapy. Whilst this may prevent the effective use of these combinations, there are existing strategies to manage haematological side-effects, such as dose interruptions, dose reductions and in appropriate patients, treat with granulocyte colony- stimulating factors to help stimulate neutrophil production [73, 74], and these could be applied in the combination setting along with close monitoring of patients. To our knowledge, this is the first synthesised analysis of the efficacy of combination PARP inhibitor and chemotherapy across multiple malignancies to demonstrate that this benefit is dependent upon the presence of a DNA repair biomarker.

## Data Availability

All data produced in the present study are available upon reasonable request to the authors.

## ACKNOWLEDGEMENTS

We thank Dr Mark Donoghoe of Stats Central, Mark Wainwright Analytical Centre, UNSW Sydney, for assistance with data extraction methods for the meta-analysis. This research was generously supported by the Dr Lee MacCormick Edwards Charitable Foundation. C.E.C. is supported by a Cancer Institute NSW Career Development Fellowship (CDF1071).

## Notes

### Competing Interest Statement

The authors have declared no competing interest.

### Author Declarations

Data from published studies was analysed as a meta-analysis, and all publications are appropriately cited in the text.

