## Supplementary data for "DNA repair biomarkers to guide usage of combined PARP inhibitors and chemotherapy: a meta-analysis and systematic review"

**Supplementary Table 1. List of 79 clinical trials investigating the efficacy of combination PARP inhibitor and chemotherapy treatment. Clinical trials were found by searching “PARP inhibitor”, “chemotherapy” and “combination” in ClinicalTrials.gov and characteristics were extracted. Note that three studies included in our meta-analysis (Liu 2018, Middleton 2015, Geyer 2022) were not identified by this search strategy. HRD: homologous recombination deficiency.**

| Title | Status | Study results available | Conditions | Interventions | Clinical trial number | Phase | Selected for HRD status | HRD status as a secondary objective | Year of initiation |
|---|---|---|---|---|---|---|---|---|---|
| BGB-290 and Temozolomide in Treating Patients with Recurrent Gliomas with IDH1/2 Mutations | Active, not recruiting | No | Glioblastoma | BGB-290, Temozolomide | NCT03914742 | I/II | Yes (IDH mutation) | - | 2019 |
| Talazoparib and Low-Dose Temozolomide in Treating Participants With Relapsed or Refractory Extensive-Stage Small Cell Lung Cancer | Active, not recruiting | No | Small Cell Lung Carcinoma | Talazoparib, Temozolomide | NCT03672773 | II | No | No | 2018 |
| ARIEL4: A Study of Rucaparib Versus Chemotherapy BRCA Mutant Ovarian, Fallopian Tube, or Primary Peritoneal Cancer Patients | Active, not recruiting | Yes | Ovarian, Fallopian Tube, Peritoneal Cancer | Chemotherapy, Rucaparib | NCT02855944 | III | Yes (*BRCA* mutation) | - | 2016 |
| Veliparib With Carboplatin and Paclitaxel and as Continuation Maintenance Therapy in Adults With Newly Diagnosed Stage III or IV, High-grade Serous, Epithelial Ovarian, Fallopian Tube, or Primary Peritoneal Cancer | Active, not recruiting | Yes | Ovarian Cancer | Veliparib, Paclitaxel, Carboplatin | NCT02470585 | III | Yes (*BRCA* mutation and HRD score) | - | 2015 |
| Testing the Addition of the Anti-Cancer Drug Talazoparib to the Combination of Carboplatin and Paclitaxel for the Treatment of Advanced Cancer | Active, not recruiting | No | Advanced Malignant Solid Neoplasm | Carboplatin, Paclitaxel, Talazoparib | NCT02317874 | I | Yes (*BRCA* mutation) | - | 2014 |
| A Phase 3 Randomized, Placebo-controlled Trial of Carboplatin and Paclitaxel With or Without Veliparib (ABT-888) in HER2-negative Metastatic or Locally Advanced Unresectable BRCA-associated Breast Cancer | Active, not recruiting | Yes | Breast Cancer | Veliparib, Carboplatin, Paclitaxel | NCT02163694 | III | Yes (*BRCA* mutation) | - | 2014 |

| | | | | | | | | | |
|---|---|---|---|---|---|---|---|---|---|
| Gemcitabine Hydrochloride and Cisplatin With or Without Veliparib or Veliparib Alone in Treating Patients With Locally Advanced or Metastatic Pancreatic Cancer | Active, not recruiting | No | Pancreatic Cancer | Cisplatin, Gemcitabine, Veliparib | NCT01585805 | II | Yes (*BRCA, PALB2* mutation) | - | 2012 |
| Veliparib, Paclitaxel, and Carboplatin in Treating Patients With Solid Tumors That Are Metastatic or Cannot Be Removed by Surgery and Liver or Kidney Dysfunction | Active, not recruiting | No | Metastatic Solid Tumours | Carboplatin, Paclitaxel, Veliparib | NCT01366144 | I | No | No | 2011 |
| Cyclophosphamide and Veliparib in Treating Patients With Locally Advanced or Metastatic Breast Cancer | Active, not recruiting | No | Breast Cancer | Cyclophosphamide, Veliparib | NCT01351909 | I | No | No | 2011 |
| Veliparib With or Without Carboplatin in Treating Patients With Stage IV Breast Cancer | Active, not recruiting | No | Breast Cancer | Carboplatin, Veliparib | NCT01149083 | II | Yes (*BRCA* mutation) | - | 2010 |
| Veliparib and Temozolomide in Treating Patients With Acute Leukemia | Active, not recruiting | No | Leukaemia | Temozolomide, Veliparib | NCT01139970 | I | No | Yes (average expression of MGMT) | 2010 |
| Study to Compare the Efficacy and Safety of Olaparib When Given in Combination With Carboplatin and Paclitaxel, Compared With Carboplatin and Paclitaxel in Patients With Advanced Ovarian Cancer | Active, not recruiting | Yes | Ovarian Cancer | Olaparib, Paclitaxel, Carboplatin | NCT01081951 | II | No | No | 2010 |
| Efficacy Study of Olaparib With Paclitaxel Versus Paclitaxel in Gastric Cancer Patients | Active, not recruiting | Yes | Gastric Cancer | Olaparib, Paclitaxel | NCT01063517 | II | Yes (ATM protein status) | - | 2010 |
| Veliparib and Topotecan Hydrochloride in Treating Patients With Solid Tumors, Relapsed or Refractory Ovarian Cancer, or Primary Peritoneal Cancer | Active, not recruiting | No | Ovarian Cancer | Topotecan Hydrochloride, Veliparib | NCT01012817 | I/II | No | Yes (*BRCA* mutation) | 2009 |
| ABT-888 and Temozolomide for Metastatic Breast Cancer and BRCA1/2 Breast Cancer | Active, not recruiting | No | Breast Cancer | ABT-888, Temozolomide | NCT01009788 | II | No | Yes (expansion cohort investigating *BRCA* mutation) | 2009 |
| Veliparib, Cyclophosphamide, and Doxorubicin Hydrochloride in Treating Patients With Metastatic or Unresectable Solid Tumors or Non-Hodgkin Lymphoma | Active, not recruiting | No | Non-Hodgkin Lymphoma, Solid Neoplasm | Cyclophosphamide, Doxorubicin Veliparib | NCT00740805 | I | No | Yes (Phase II cohort enrols *BRCA* mutated patients) | 2008 |
| Veliparib and Irinotecan Hydrochloride in Treating Patients With Cancer That Is Metastatic or Cannot Be Removed by Surgery | Active, not recruiting | No | Metastatic Solid Neoplasms | Irinotecan Hydrochloride, Veliparib | NCT00576654 | I | No | Yes (expansion cohort investigates | 2007 |

| | | | | | | | | | |
|---|---|---|---|---|---|---|---|---|---|
| | | | | | | | | *BRCA* mutation) | |
| Study to Assess the Safety and Tolerability of a PARP Inhibitor in Combination With Carboplatin and/or Paclitaxel | Active, not recruiting | No | Breast Cancer, Ovarian Cancer | AZD2281, Carboplatin, Paclitaxel | NCT00516724 | I | No | No | 2007 |
| Rucaparib and Irinotecan in Cancers With Mutations in DNA Repair | Completed | No | Solid Neoplasms | Rucaparib, Irinotecan | NCT03318445 | I | Yes (DNA repair defects) | - | 2017 |
| Study of Gemcitabine, Carboplatin and VELIPARIB (ABT-888) in Refractory Testicular Germ Cell Cancer | Completed | No | Testicular Cancer | Veliparib, Gemcitabine, Carboplatin | NCT02860819 | II | No | No | 2016 |
| Veliparib in Combination With Carboplatin And Weekly Paclitaxel in Japanese Subjects With Ovarian Cancer | Completed | No | Ovarian Cancer | Veliparib, Carboplatin, Paclitaxel | NCT02483104 | I | No | No | 2015 |
| Talazoparib Plus Irinotecan With or Without Temozolomide in Children With Refractory or Recurrent Solid Malignancies | Completed | No | Childhood Solid Tumors | Talazoparib, Irinotecan, Temozolomide, Filgrastim, Peg-filgrastim | NCT02392793 | I | No | No | 2015 |
| Dose Escalation and Double-blind Study of Veliparib in Combination With Carboplatin and Etoposide in Treatment-naive Extensive Stage Disease Small Cell Lung Cancer | Completed | Yes | Small Cell Lung Cancer | Veliparib, Carboplatin, Etoposide | NCT02289690 | I/II | No | No | 2014 |
| Study Comparing Veliparib Plus Carboplatin and Paclitaxel Versus Investigator's Choice of Standard Chemotherapy in Adults Receiving First Cytotoxic Chemotherapy for Metastatic or Advanced Non-Squamous Non-Small Cell Lung Cancer (NSCLC) and Who Are Current or Former Smokers | Completed | Yes | Non-squamous Non-small Cell Lung Cancer | Paclitaxel, Carboplatin, Cisplatin, Veliparib, Pemetrexed | NCT02264990 | III | No | No | 2014 |
| Study Comparing Veliparib Plus Carboplatin and Paclitaxel Versus Placebo Plus Carboplatin and Paclitaxel in Previously Untreated Advanced or Metastatic Squamous Non-Small Cell Lung Cancer | Completed | No | Squamous Non-Small Cell Lung Cancer | Carboplatin, Veliparib, Paclitaxel | NCT02106546 | III | No | No | 2014 |
| ESP1/SARC025 Global Collaboration: A Phase I Study of a Combination of the PARP Inhibitor, Niraparib and Temozolomide and/or Irinotecan Patients With Previously Treated, Incurable Ewing Sarcoma | Completed | No | Ewing Sarcoma | Niraparib, Temozolomide, Irinotecan | NCT02044120 | I | No | No | 2014 |

| | | | | | | | | | |
|---|---|---|---|---|---|---|---|---|---|
| A Study Evaluating Veliparib as a Single Agent or in Combination With Chemotherapy in Subjects With Solid Tumors | Completed | No | Solid Tumors | Veliparib, Carboplatin, Paclitaxel, FOLFIRI | NCT02033551 | I | Yes (DNA repair defects) | - | 2014 |
| Veliparib and Floxuridine in Treating Patients With Metastatic Epithelial Ovarian, Primary Peritoneal Cavity, or Fallopian Tube Cancer | Completed | No | Ovarian, Fallopian Tube, Peritoneal Cancer | Floxuridine, Veliparib | NCT01749397 | I | No | Yes (assesses presence of HRD mutations) | 2012 |
| Cisplatin and Etoposide With or Without Veliparib in Treating Patients With Extensive Stage Small Cell Lung Cancer | Completed | Yes | Small Cell Lung Carcinoma | Cisplatin, Etoposide, Veliparib | NCT01642251 | II | No | No | 2012 |
| Temozolomide With or Without Veliparib in Treating Patients With Relapsed or Refractory Small Cell Lung Cancer | Completed | Yes | Small Cell Lung Carcinoma | Temozolomide, Veliparib | NCT01638546 | II | No | Yes (DNA damage markers as other outcome measures) | 2012 |
| An Open-Label, Multicenter, Phase 1/2 Study of Poly(ADP-Ribose) Polymerase (PARP) Inhibitor E7449 as Single Agent in Subjects With Advanced Solid Tumors or With B-cell Malignancies and in Combination With Temozolomide (TMZ) or With Carboplatin and Paclitaxel in Subjects With Advanced Solid Tumors | Completed | No | Solid Tumors | E7449, Temozolomide, Carboplatin, Paclitaxel | NCT01618136 | I/II | No | Yes (Phase 2 enrols ATM-deficient patients) | 2012 |
| A Clinical Study Conducted in Multiple Centers Comparing Veliparib in Combination With Carboplatin and Paclitaxel Versus a Placebo in Combination With Carboplatin and Paclitaxel in Patients With Advanced Non-small Cell Lung Cancer | Completed | No | Non-Small Cell Lung Cancer | Veliparib, Carboplatin, Paclitaxel | NCT01560104 | II | No | No | 2012 |
| Study Evaluating Efficacy And Tolerability Of Veliparib in Combination With Temozolomide (TMZ) or In Combination With Carboplatin and Paclitaxel Versus Placebo in Participants With Breast Cancer Gene (BRCA)1 and BRCA2 Mutation and Metastatic Breast Cancer | Completed | Yes | Breast Cancer | Veliparib, Carboplatin, Temozolomide, Paclitaxel | NCT01506609 | II | Yes (*BRCA* mutation) | - | 2012 |
| ABT-888 Plus Metronomic Cyclophosphamide to Treat Cancer | Completed | No | Neoplasms, Lymphoma | ABT-888, Cyclophosphamide | NCT01445522 | I | No | No | 2011 |
| AZD2281 Plus Carboplatin to Treat Breast and Ovarian Cancer | Completed | No | Breast Cancer, Ovarian Cancer | AZ2281, Carboplatin | NCT01445418 | I | Yes (*BRCA* mutation) | - | 2011 |

| | | | | | | | | | |
|---|---|---|---|---|---|---|---|---|---|
| Olaparib and Temozolomide in Treating Patients With Relapsed Glioblastoma | Completed | No | Brain, Central Nervous System Tumors | Olaparib, Temozolomide | NCT01390571 | I | No | Yes (secondary analysis of DNA repair genes) | 2011 |
| Study of CEP-9722 in Combination With Gemcitabine and Cisplatin in Patients With Advanced Solid Tumors or Mantle Cell Lymphoma | Completed | No | Solid Tumors, Mantle Cell Lymphoma | CEP-9722, Gemcitabine, Cisplatin | NCT01345357 | I | No | No | 2011 |
| Phase II ABT-888 With Cyclophosphamide | Completed | Yes | Ovarian, Peritoneal, Breast, Fallopian Tube Cancer | ABT-888, Cyclophosphamide | NCT01306032 | II | No | Yes (Group 1 enrols *BRCA*-positive patients) | 2011 |
| Paclitaxel, Cisplatin, and Veliparib in Treating Patients With Advanced, Persistent, or Recurrent Cervical Cancer | Completed | No | Cervical Cancer | Cisplatin, Paclitaxel, Veliparib | NCT01281852 | I | No | Yes (secondary analysis of loss of Fanconi Anemia formation) | 2011 |
| Veliparib in Combination With Carboplatin and Paclitaxel in Treating Patients With Locally Advanced or Metastatic Solid Tumors | Completed | No | Solid Tumours | Carboplatin, Paclitaxel, Veliparib | NCT01281150 | I | No | Yes (secondary analysis of HRD genes) | 2011 |
| Veliparib and Carboplatin in Treating Patients With HER2-Negative Metastatic Breast Cancer | Completed | No | Breast Cancer | Carboplatin, Fluorothymidine F-18, Veliparib | NCT01251874 | I | No | Yes (secondary analysis of DNA repair genes e.g. *BRCA* and Fanconi Anemia pathway genes) | 2010 |
| Olaparib in Combination With Carboplatin for Refractory or Recurrent Womens Cancers | Completed | No | Ovarian, Breast, Peritoneal, Fallopian Tube, Endometrial Cancer | Carboplatin, Olaparib | NCT01237067 | I | No | No | 2010 |
| ABT-888 and Gemcitabine Hydrochloride in Treating Patients With Advanced Solid Tumors | Completed | No | Solid Tumours | Gemcitabine Hydrochloride, Veliparib | NCT01154426 | I | Yes (*BRCA* mutation) | - | 2010 |
| Veliparib and Pegylated Liposomal Doxorubicin Hydrochloride in Treating Patients With Recurrent Ovarian Cancer, Fallopian Tube Cancer, or Primary Peritoneal Cancer or Metastatic Breast Cancer | Completed | No | Ovarian, Breast, Peritoneal, Fallopian Tube Cancer | Pegylated Liposomal Doxorubicin Hydrochloride, Veliparib | NCT01145430 | I | No | Yes (secondary analysis of *BRCA* mutation) | 2010 |
| Study of Poly (ADP-Ribose) Polymerase (PARP) Inhibitor E7016 in Combination With Temozolomide in Subjects With Advanced Solid Tumors | Completed | No | Solid Tumors | E7016, Temozolomide | NCT01127178 | I | No | No | 2010 |
| A Study of ABT-888 in Combination With Carboplatin and Gemcitabine in | Completed | No | Solid Tumors | Veliparib, Carboplatin, Gemcitabine | NCT01063816 | I | No | No | 2010 |

| | | | | | | | | | |
|---|---|---|---|---|---|---|---|---|---|
| Subjects With Advanced Solid Tumors | | | | | | | | | |
| A Study of ABT-888 in Combination With Temozolomide for Colorectal Cancer | Completed | Yes | Colorectal Cancer | Temozolomide, ABT-888 | NCT01051596 | II | No | No | 2010 |
| Veliparib With or Without Mitomycin C in Treating Patients With Metastatic, Unresectable, or Recurrent Solid Tumors | Completed | No | Solid Neoplasm | Mitomycin,Veliparib | NCT01017640 | I | Yes (deficient Fanconi Anemia pathway) | - | 2009 |
| A Study Of Poly (ADP-Ribose) Polymerase Inhibitor PF-01367338 In Combination With Several Chemotherapeutic Regimens | Completed | No | Solid Tumours | PF-01367338, Carboplatin | NCT01009190 | I | No | No | 2009 |
| A Phase I Study of ABT-888, an Oral Inhibitor of Poly(ADP-ribose) Polymerase and Temozolomide in Children With Recurrent/Refractory CNS Tumors | Completed | No | Central Nervous System Tumours | Temozolomide, ABT-888 | NCT00994071 | I | No | No | 2009 |
| AZD2281 and Cisplatin Plus Gemcitabine to Treat Solid Tumor Cancers | Completed | No | Neoplasms | AZD 2281, Cisplatin, Gemcitabine | NCT00678132 | I | No | No | 2008 |
| Safety Study of ABT-888 Plus Topotecan Hydrochloride to Treat Patients With Solid Tumors and Lymphomas | Completed | No | Solid Tumors, Lymphomas | ABT-888, Topotecan | NCT00553189 | I | No | No | 2007 |
| Veliparib, Carboplatin, and Paclitaxel in Treating Patients With Advanced Solid Cancer | Completed | No | Solid Tumours | Carboplatin, Paclitaxel, Veliparib | NCT00535119 | I | No | Yes (Phase II cohort investigates *BRCA* mutation) | 2007 |
| A Phase I Study of ABT-888 in Combination With Temozolomide in Cancer Patients | Completed | No | Solid Tumours | ABT-888, Temozolomide | NCT00526617 | I | Yes (*BRCA* mutation) | - | 2007 |
| Study to Assess the Safety and Tolerability of a PARP Inhibitor in Combination With Topotecan | Completed | No | Solid Tumors | AZD2281, Topotecan | NCT00516438 | I | No | No | 2007 |
| Study to Assess the Safety & Tolerability of a PARP Inhibitor in Combination With Gemcitabine in Pancreatic Cancer | Completed | No | Pancreatic Neoplasms | AZD2281, Gemcitabine | NCT00515866 | I | No | No | 2007 |
| Testing Olaparib and Temozolomide Versus the Usual Treatment for Uterine Leiomyosarcoma After Chemotherapy Has Stopped Working | Not yet recruiting | No | Uterine Leiomyosarcoma | Olaparib, Pazopanib, Temozolomide, Trabectedin | NCT05432791 | II/III | No | Yes (exploratory analysis of HRD genes) | 2022 |
| EP0057 in Combination With Olaparib in Relapsed Advanced Gastric Cancer and Small Cell Lung Cancer | Not yet recruiting | No | Gastric, Small-cell Lung Cancer | EP0057, Olaparib | NCT05411679 | II | No | Yes (Arm 1 enrols ATM-negative patients and | 2022 |

| | | | | | | | | | |
|---|---|---|---|---|---|---|---|---|---|
| | | | | | | | | secondary analysis investigates HRD status) | |
| Study to Assess the Safety, Tolerability of JPI-547 in Combination With Modified FOLFIRINOX or Gemcitabine-nab-paclitaxel in Patients With Locally Advanced and Metastatic Pancreatic Cancer | Not yet recruiting | No | Pancreatic Ductal Adenocarcinoma | JPI-547, FOLFIRINOX, Gemcitabine-nab-paclitaxel | NCT05257993 | I | No | No | 2022 |
| Temozolomide Monotherapy or in Combination With Olaparib in Patients With Triple Negative Breast Cancer (TNBC) | Not yet recruiting | No | Breast Cancer | Temozolomide, Olaparib | NCT05128734 | II | Yes (MGMT promoter methylated) | - | 2021 |
| Relapsed Pediatric AML to Determine the Safety and Efficacy of the PARP Inhibitor Talazoparib in Combination With Conventional Chemotherapy (POE22-01) (PARPAML) | Recruiting | No | Acute Myeloid Leukemia | Talazoparib, Topotecan, Gemcitabine | NCT05101551 | I | No | No | 2021 |
| Testing the Combination of Anti-Cancer Drugs Talazoparib and Temozolomide in Patients >= 18 Years Old With Advanced Stage Rare Cancers, RARE 2 Study | Recruiting | No | Adrenal Gland Pheochromocytoma, Hematopoietic and Lymphoid System Neoplasm, Malignant Solid Neoplasm, Paraganglioma | Talazoparib, Temozolomide | NCT05142241 | II | No | No | 2021 |
| Pamiparib and Temozolomide for the Treatment of Hereditary Leiomyomatosis and Renal Cell Cancer | Recruiting | No | Leiomyomatosis, Renal Cell Cancer | Pamiparib, Temozolomide | NCT04603365 | II | No | Potential (secondary analysis of genomic mutational signature via whole genome sequencing) | 2020 |
| NUVOLA TRIAL Open-label Multicentre Study | Recruiting | No | High Grade Serous Ovarian Cancer | Olaparib, Paclitaxel, Carboplatin | NCT04261465 | II | Yes (*BRCA* mutation) | - | 2020 |
| A Study of Fluzoparib in Combination With mFOLFIRINOX in Patients With Advanced Pancreatic Cancer | Recruiting | No | Advanced Pancreatic Cancer | Fluzoparib,\| mFOLFIRINOX | NCT04228601 | I/II | Yes (*BRCA, PALB2* mutation) | - | 2020 |
| PLX038 (PEGylated SN38) and Rucaparib in Solid Tumors and Small Cell Cancers | Recruiting | No | Small Cell Lung Cancer, Extra-Pulmonary Small Cell Carcinomas | PLX038, Rucaparib | NCT04209595 | I/II | No | No | 2019 |
| BGB-290 and Temozolomide in Treating Isocitrate Dehydrogenase (IDH)1/2-Mutant Grade I-IV Gliomas | Recruiting | No | Glioma | BGB-290, Temozolomide | NCT03749187 | I | Yes (*IDH1/2* mutation) | - | 2018 |

| | | | | | | | | | |
|---|---|---|---|---|---|---|---|---|---|
| Niraparib Plus Carboplatin in Patients With Homologous Recombination Deficient Advanced Solid Tumor Malignancies | Recruiting | No | Solid Tumor | Niraparib, Carboplatin | NCT03209401 | I | Yes (DNA repair defects) | - | 2017 |
| Platinum and Polyadenosine 5'Diphosphoribose Polymerisation (PARP) Inhibitor for Neoadjuvant Treatment of Triple Negative Breast Cancer (TNBC) and/or Germline BRCA (gBRCA) Positive Breast Cancer | Recruiting | No | Breast Cancer | Olaparib, Paclitaxel and Carboplatin | NCT03150576 | II/III | No (*BRCA* mutation inclusion criteria but not necessary for enrolment) | No | 2017 |
| Phase I Study of Olaparib and Temozolomide for Ewings Sarcoma or Rhabdoomyosarcoma | Recruiting | No | Ewing Sarcoma, Rhabdomyosarcoma | Olaparib, Temozolomide, Irinotecan | NCT01858168 | I | No | No | 2013 |
| A Study Evaluating the Efficacy and Tolerability of Veliparib in Combination With Paclitaxel/Carboplatin-Based Chemoradiotherapy Followed by Veliparib and Paclitaxel/Carboplatin Consolidation in Adults With Stage III Non-Small Cell Lung Cancer (NSCLC) | Terminated | Yes | Non-small Cell Lung Cancer Stage | Paclitaxel, Veliparib, Carboplatin | NCT02412371 | I/II | No | No | 2015 |
| ABT-888 and Temozolomide for Liver Cancer | Terminated | Yes | Hepatocellular Carcinoma | Temozolomide, ABT-888 | NCT01205828 | II | No | Yes (secondary analysis of *BRCA1/2* mutation and DNA repair genes) | 2010 |
| A Study of Fluzoparib Given in Combination With Apatinib and Paclitaxel in Gastric Cancer Patients | Unknown status | No | Gastric Cancer | Fluzoparib, Apatinib, Paclitaxel | NCT03026881 | I | No | No | 2017 |
| Combination of Carboplatin, Eribulin and Veliparib in Stage IV Cancer Patients | Withdrawn | No | Breast, Ovarian Cancer | Carboplatin, Eribulin, Veliparib | NCT03032614 | II | Yes (*BRCA1/2* mutation, PTEN deficiency, high HRD score) | - | 2017 |
| A Study Evaluating Talazoparib in Relapsed Ovarian, Fallopian Tube, and Peritoneal Cancer | Withdrawn | No | Ovarian Cancer | Talazoparib, Temozolomide | NCT02836028 | II | Yes (*BRCA1/2* mutation, high HRD score) | - | 2016 |
| ABT-888, Carboplatin, and Paclitaxel for Cancer With Liver or Kidney Problems | Withdrawn | No | Neoplasms | ABT-888, Carboplatin, Paclitaxel | NCT01419548 | I | No | No | 2011 |
| Topotecan Hydrochloride and Carboplatin With or Without Veliparib in Treating Advanced Myeloproliferative Disorders and Acute Myeloid Leukemia or Chronic Myelomonocytic Leukemia | Active, not recruiting | No | Leukemia | Veliparib, Topotecan Hydrochloride, Carboplatin | NCT03289910 | II | No | No | 2017 |

| | | | | | | | | | |
|---|---|---|---|---|---|---|---|---|---|
| EP0057 in Combination With Olaparib in Advanced Ovarian Cancer | Active, not recruiting | No | Ovarian cancer | Olaparib, EP007, Camptothecin | NCT04669002 | II | No (Phase 2A cohort, *BRCA* and HRD mutational status does not need to be known) | No | 2020 |

| Study ID | Outcome | D1 | D2 | D3 | D4 | D5 | Overall |
|---|---|---|---|---|---|---|---|
| Byers 2021 | PFS | + | + | + | + | + | + |
| Coleman 2019 | PFS | + | + | + | + | + | + |
| Geyer 2020 | PFS | + | + | + | + | + | + |
| Kummar 2015 | PFS | ! | + | + | ! | + | ! |
| Liu 2018 | PFS | + | + | + | + | + | + |
| Middleton 2015 | PFS | + | + | + | + | + | + |
| Oza 2015 | PFS | + | + | + | + | + | + |
| Pietanza 2018 | PFS | + | + | + | + | + | + |
| Sharma 2020 | PFS | ! | ! | + | ! | ! | ! |

+ Low risk
! Some concerns
- High risk

D1 Randomisation process
D2 Deviations from the intended interventions
D3 Missing outcome data
D4 Measurement of the outcome
D5 Selection of the reported result

**Supplementary Figure 1. Risk-of-bias assessment of studies in primary progression-free survival analyses.**

| Study ID | Outcome | D1 | D2 | D3 | D4 | D5 | Overall |
|---|---|---|---|---|---|---|---|
| Bang 2017 | AE | + | + | + | + | + | + |
| Byers 2021 | AE | + | + | + | + | + | + |
| Coleman 2019 | AE | + | + | + | + | + | + |
| Dieras 2020 | AE | + | + | + | + | + | + |
| Govindan 2022 | AE | + | + | + | ! | + | ! |
| Han 2018 | AE | + | + | + | ! | + | ! |
| Oza 2015 | AE | + | + | + | + | + | + |
| Ramalingam 2021 | AE | + | + | + | + | + | + |

\+ Low risk
! Some concerns
\- High risk

D1 Randomisation process
D2 Deviations from the intended interventions
D3 Missing outcome data
D4 Measurement of the outcome
D5 Selection of the reported result

**Supplementary Figure 2. Risk-of-bias assessment of studies in secondary adverse effects analyses**

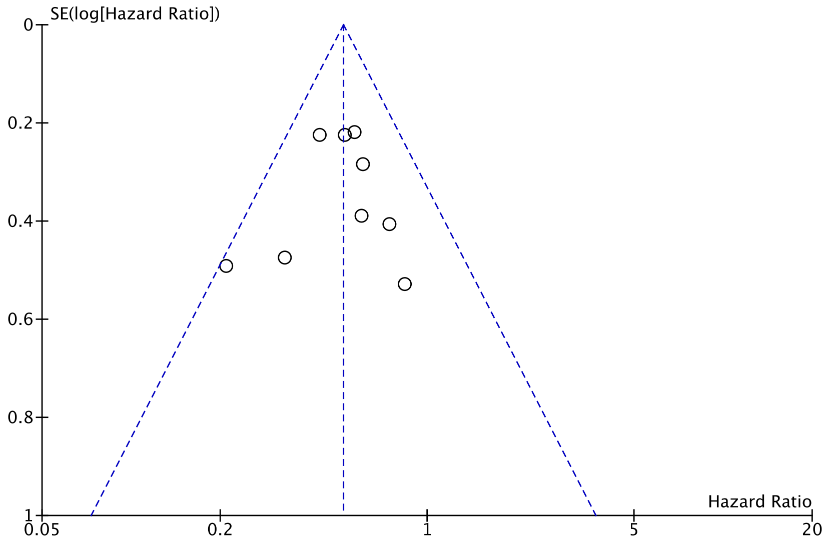


**Supplementary Figure 3. Funnel plot investigating publication bias for DNA repair deficient progression-free survival endpoint.**

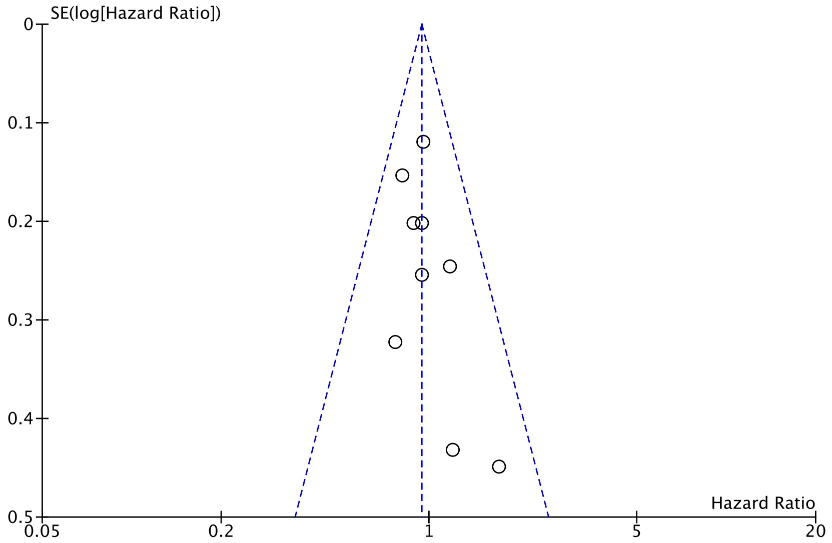


**Supplementary Figure 4. Funnel plot investigating publication bias for DNA repair competent progression-free survival endpoint.**

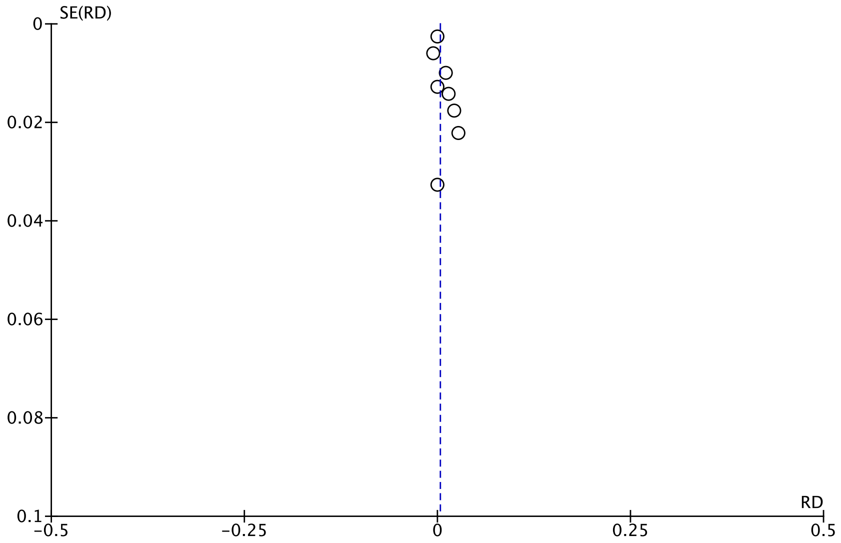


**Supplementary Figure 5. Funnel plot investigating publication bias for all adverse effects endpoint. SE(RD) is standard error of risk difference.**

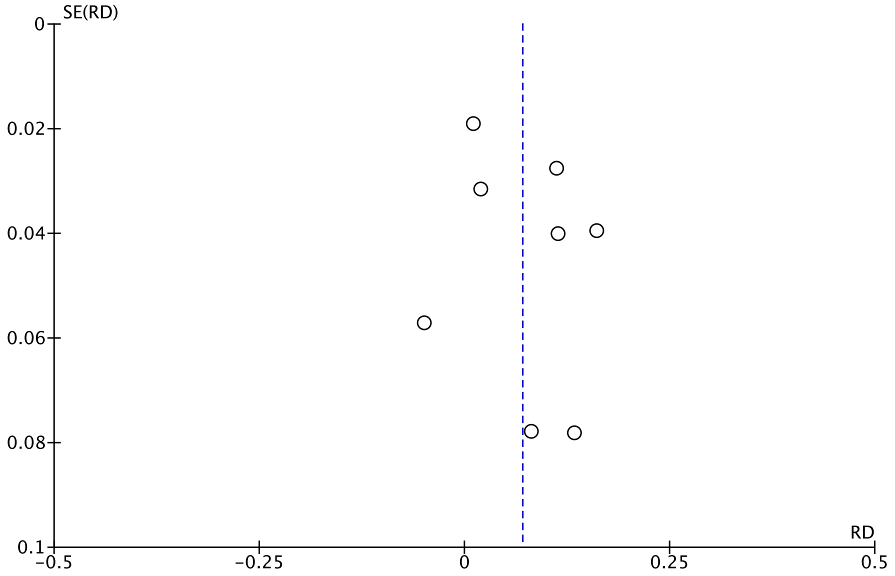


**Supplementary Figure 6. Funnel plot investigating publication bias for grade ¾ adverse effects endpoint. SE(RD) is standard error of risk difference.**

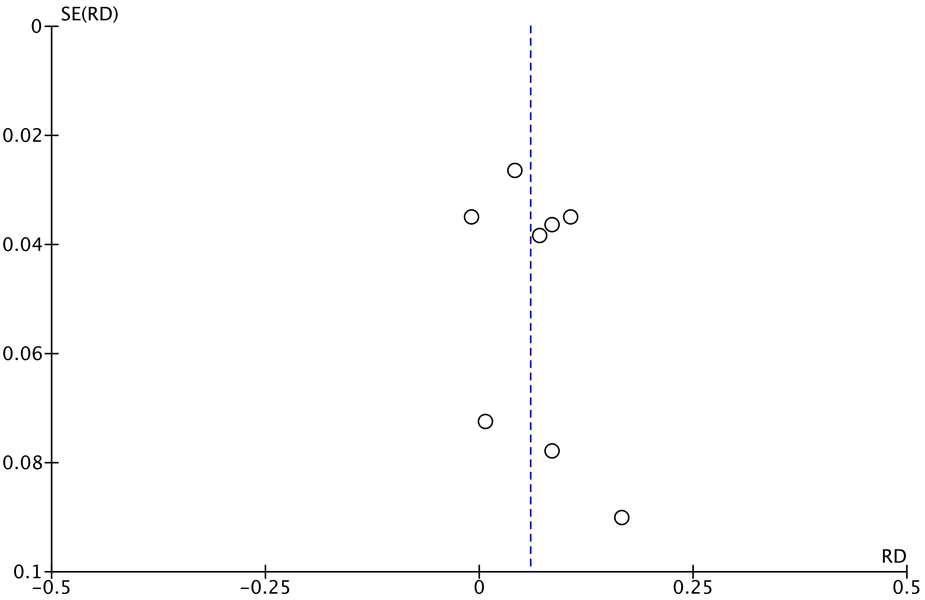


**Supplementary Figure 7. Funnel plot investigating publication bias for hematological (neutropenia) grade ¾ adverse effects endpoint. SE(RD) is standard error of risk difference.**
